# Estimating the time-varying reproduction number for COVID-19 in South Africa during the first four waves using multiple measures of incidence for public and private sectors across four waves

**DOI:** 10.1101/2022.07.22.22277932

**Authors:** Jeremy Bingham, Stefano Tempia, Harry Moultrie, Cecile Viboud, Waasila Jassat, Cheryl Cohen, Juliet R.C. Pulliam

**Affiliations:** South African DSI-NRF Centre of Excellence in Epidemiological Modelling and Analysis (SACEMA), Stellenbosch University, Stellenbosch, South Africa; Centre for Respiratory Diseases and Meningitis, National Institute for Communicable Diseases of the National Health Laboratory Service, Johannesburg, South Africa; School of Public Health, University of the Witwatersrand, Johannesburg, South Africa; Centre for Tuberculosis, National Institute for Communicable Diseases, Division of the National Health Laboratory Service, Johannesburg, South Africa; School of Pathology, Faculty of Health Sciences, University of the Witwatersrand, Johannesburg, South Africa; Fogarty International Center, NIH, Bethesda, MD, USA; Division of Public Health Surveillance and Response, National Institute for Communicable Diseases, National Health Laboratory Service, Johannesburg, South Africa; Right to Care, Pretoria, South Africa

## Abstract

**Objectives:** We aimed to quantify transmission trends in South Africa during the first four waves of the COVID-19 pandemic using estimates of the time-varying reproduction number (R) and to compare the robustness of R estimates based on three different data sources and using data from public and private sector service providers.

**Methods:** We estimated R from March 2020 through April 2022, nationally and by province, based on time series of rt-PCR-confirmed cases, hospitalizations, and hospital-associated deaths, using a method which models daily incidence as a weighted sum of past incidence. We also estimated R separately using public and private sector data.

**Results:** Nationally, the maximum case-based R following the introduction of lockdown measures was 1.55 (CI: 1.43-1.66), 1.56 (CI: 1.47-1.64), 1.46 (CI: 1.38-1.53) and 3.33 (CI: 2.84-3.97) during the first (Wuhan-Hu), second (Beta), third (Delta), and fourth (Omicron) waves respectively. Estimates based on the three data sources (cases, hospitalisations, deaths) were generally similar during the first three waves but case-based estimates were higher during the fourth wave. Public and private sector R estimates were generally similar except during the initial lockdowns and in case-based estimates during the fourth wave.

**Discussion:** Agreement between R estimates using different data sources during the first three waves suggests that data from any of these sources could be used in the early stages of a future pandemic. High R estimates for Omicron relative to earlier waves is interesting given a high level of exposure pre-Omicron. The agreement between public and private sector R estimates highlights the fact that clients of the public and private sectors did not experience two separate epidemics, except perhaps to a limited extent during the strictest lockdowns in the first wave.

## Introduction

As of April 2022, South Africa had experienced four waves of COVID-19, following the first reported case in early March 2020. In March 2020, while case numbers were still low, the South African government declared a national state of emergency and introduced strict lockdown legislation, with five lockdown “levels” [1–3]. Lockdown started at level five, which forced the temporary closure of all non-essential businesses, the closure of international and inter-provincial borders, an alcohol and tobacco prohibition, and a ban on leaving one’s home except to access essential services. Lockdown restrictions relaxed, due to economic pressure, early during the onset of the first wave, and were re-introduced in adjusted forms during the second and third waves. The first wave was associated with primarily wild type SARS-COV-2, the second wave with the Beta variant (B.1.351), the third wave with the Delta variant (B.1.617.2), and the fourth wave with the Omicron variant (B.1.1.529.1) [4]. During July 2021, civil unrest caused disruptions to laboratory and healthcare services in KwaZulu-Natal and Gauteng provinces.

In South Africa, roughly 17% of residents have access to private healthcare through health insurance, while the remaining 83% rely primarily on the public healthcare system [5]. Clients of the private sector have a higher mean income and tend to live in areas with lower population density [6,7]; in addition, people with lower socioeconomic status are likely to be at higher risk of SARS-COV-2 infection [7–10]. As such, transmission patterns may be expected to differ between clients of private vs public sector healthcare providers, due to the difficulty of adhering to lockdown measures in higher-density areas [11].

The time-varying reproduction number R is the expected number of secondary cases caused by a single infected individual at a given point in time, assuming that conditions remain constant for the duration of the infectious period. R reflects inherent pathogen properties combined with environmental and social conditions such as population immunity, non-pharmaceutical interventions (NPIs), perceptions of disease, and access to healthcare. Reproduction number estimates are used to track transmission trends, assess the impacts of interventions, and parameterize epidemic models [12,13].

Several studies featuring R estimates for South Africa have been published since the start of the COVID-19 pandemic. However, aside from the regular reports we released via the National Institute for Communicable Diseases (NICD) [14], the studies we identified relied on publicly reported time series data, which do not include symptom onset dates and which may be affected by backfilling and additional reporting delays, and used international estimates for the generation interval [1,2,15–21]; most studies also use a single measure of incidence and do not cover all of the first four waves. While currently-available evidence suggests that clients of the public sector experienced higher levels of transmission during the first two waves [7,22], we did not identify any studies comparing reproduction number estimates in different income groups, or between healthcare sectors. R estimates are typically based on time series data of cases or deaths, though any measure representing an approximately constant proportion of total incidence may be used, in conjunction with data from which to estimate the generation interval [23]. For example, time series of deaths are sometimes used for reproduction number estimation, particularly in situations where the consistency of testing systems is called into question, as deaths are thought to be more systematically assessed and recorded than other measures of incidence.

We generated R estimates for COVID-19 in South Africa during the first four waves of the epidemic. We examined whether R estimates differed according to data source (comparing the daily incidence of laboratory-confirmed COVID-19 cases, hospitalizations, and in-hospital deaths), and compared transmission patterns between the public and private sectors.

## Methods

### Data

Our analyses were based on two primary data sets maintained by the South African NICD, from the first confirmed case on March 5^th^ 2020 through April 28^th^ 2022. Data on laboratory-confirmed cases were obtained from the national Notifiable Medical Conditions Surveillance System (NMC-SS) line list, to which all pathology laboratories are legally required to report positive COVID-19 test results [24]. Data on laboratory-confirmed cases includes both primary infections as well as suspected reinfections. Positive tests were classified as associated with a reinfection if more than 90 days had elapsed since the most recent positive test for the same patient [25]. Data on laboratory-confirmed cases were filtered to include only cases confirmed via reverse-transcription polymerase chain reaction (rt-PCR) testing, due to substantial numbers of incorrectly entered reference dates for antigen tests. Data on hospital admissions and hospital-associated deaths were obtained from the national DATCOV dataset, to which all private (262) and public (407) hospitals report confirmed COVID-19-positive admissions and deaths in hospital [26]. The generation interval distribution was approximated using a gamma distribution fit to data from PHIRST-C, a community cohort study of COVID-19 transmission (mean = 6.63 days, si_mean_=0.51 days; si = 3.28 days, si_si_ = 0.27 days; see R estimation subsection below) [27]. Dates of symptom onset were available for 55% of hospitalized cases and 57% of in hospital deaths. 31 cases (0.007%) in the DATCOV database were missing both admission date and date of symptom onset and were excluded from the analyses based on admissions and deaths. The reinfections line list was linked with DATCOV to obtain dates of symptom onset for 5.6% of rt-PCR-confirmed cases (obtained through the DATCOV database).

### Imputation

Missing onset dates in the source datasets were imputed using the *multiple imputation chained equations* technique, as implemented in the R packages *mice* [28] and *countimp* [29]. The imputation procedure recursively estimates individual-level delays between symptom onset and hospital admission (for hospitalizations and deaths), or between symptom onset and date of reported case confirmation (for rt-PCR-confirmed cases). In each estimation step, a multivariate negative binomial model was fitted to predict delays based on 6 (lab-confirmed cases) or 7 (hospitalizations and deaths) other variables and used to predict delay values which were incurred but not reported. Poisson-distributed generalized linear models were fit to explore the relationships between the known delay values and individual predictor variables, in order to select the most relevant predictors. The variables used for imputations in the two line lists were: health sector where laboratory testing/hospital admission occurred (public or private), age group (in ten-year intervals), month of case report/hospital admission, case outcome (for admissions), day of hospital admission (for admissions), province (for rt-PCR-confirmed cases), and district (for admissions).

### Adjustment for right-censoring

Two mechanisms exist by which the data are right-censored. The first occurs because some case confirmations, hospital admissions, and in-hospital deaths correspond to dates of symptom onset which fall within the date range of the data, despite the events (case confirmation, hospital admission, or death) occurring outside the date range of the data. The second cause of right-censoring in the data comes from case confirmations, hospital admissions, and in-hospital deaths which occur within the date range of the data, but which are not yet included in the dataset.

The first source of right-censoring was accounted for by inflating the end of the time series according to the distributions of delays from symptom onset to reporting of test results, hospital admission, and death [23]. Specifically, counts for each day were divided by the proportion of the appropriate delay distribution with delay larger than the difference between the day in question and the last date for which test results, hospital admissions, or deaths were reported. The second source of right-censoring was more difficult to rigorously adjust for, and was mitigated by truncating the last 3, 7, and 7 days (respectively) of the time series.

### R estimation

Daily time series of rt-PCR-confirmed COVID-19 cases, hospitalizations, and deaths, by dates of symptom onset, were computed using the imputed line lists and adjusted for right-censoring. R was estimated using the method described by Thompson *et al*. and implemented in the R package EpiEstim [23], identified in a recent review of R estimation techniques as a suitable method given the data available during this study [30]. The method assumes that current incidence results from the combined transmissibility of recently infected individuals, and can thus be calculated based on recent incidence values [12,23]. Specifically, the incidence *I*_*t*_ (in time step *t*) is assumed to be Poisson distributed, with mean

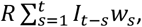

where *R* is the time-varying (instantaneous) reproduction number at time step *t*, and *w*_*s*_ is the relative infectiousness of an individual *s* time steps following infection, assumed to be constant with respect to *t*. Furthermore, R is assumed to be constant over some time window *τ*, the choice of which involves a tradeoff between the width of resulting credible intervals and the sensitivity of the method to rapid changes in *R*. We estimated R using 7, 14, and 21 day sliding windows; estimates using 7-day sliding windows are presented in the main text. The likelihoods of hypothetical values for R are calculated, resulting in a gamma-distributed Bayesian posterior for R. The infectiousness profile *w*_*s*_ was estimated using a gamma-distributed generation interval fit to data from the PHIRST-C community cohort study [27] (μ_SI_ = 6.63 days, σ_μ-SI_ =0.51 days; σ_SI_ = 3.28 days, σ_σ-SI_ = 0.27 days). In order to accommodate uncertainty in *w*_*s*_, n_1_ pairs of (μ_SI_, σ_SI_) are sampled from truncated normal distributions (with σ_SI_ < μ_SI_). From each of these n_1_ pairs, n_2_ posterior R estimates are generated, resulting in n_1_ x n_2_ R estimates, per imputation, for each time window. We generated R estimates using n_1_ = n_2_ = 25, resulting in 625 R estimates per time window per imputation. Several parameter combinations were explored to determine suitable values for n_1_ and n_2_ (see supplementary materials section 5). No explicit smoothing of time series or R estimates was performed. The R estimation procedure was performed on each of 25 imputed datasets. We present median values, between imputations, of the median R estimates and 95% credible intervals (CI) arising from the 625 individual R estimates per estimation window.

R was estimated based on rt-PCR-confirmed cases (R_cases_), hospital admissions (R_admissions_) and hospital-associated deaths (R_deaths_). R was also estimated separately using public and private sector data; rt-PCR-confirmed cases were classified according to the sector of the laboratory where samples were processed, while hospitalized cases and in-hospital deaths were classified according to the sector of the hospital where admission took place. This distinction is relevant because some patients who were admitted to public hospitals paid out of pocket to access testing services from private laboratories. Laboratory and healthcare service providers were disrupted during a period of civil unrest in KwaZulu-Natal and Gauteng provinces. We indicate the period of unrest, between 10 and 19 July 2021, and provide a simplistic indicator of its effects on R estimates by adding a shaded area to the graph, with opacity equal to the proportion of the backwards-looking generation time which occurs during or before the unrest, as this is the weight applied to past incidence values when estimating R.

## Results

### National and provincial R estimates using timeseries of cases

Nationally, R based on cases (R_cases_) dropped sharply following the closure of borders and schools in mid-March 2020 (figure 1), followed by an increase in mid-April. During levels five and four lockdown, R_cases_ fluctuated around 1.25, with large credible intervals (table 1 and figure 1). R_cases_ remained steady through the level three lockdown in June 2020, with values between 1 and 1.5, then began to decrease in late June; the maximum R_cases_ in the first wave (after April 1^st^ 2020) was 1.55 (CI: 1.43-1.66). R_cases_ crossed below 1 in mid-July and continued to decrease through early August. R_cases_ increased gradually during level two lockdown and through most of level one lockdown, then began to decrease in the first half of December while incidence in the second wave was still increasing; maximum R_casess_ in the second wave was 1.56 (CI: 1.47-1.64). The gradual decrease continued until the first week of January 2021, approximately one week after the introduction of adjusted level three lockdown, when R_cases_ declined sharply, reaching a value of 0.56 (CI: 0.53-0.60) by the end of January. R_cases_ then began to increase, in a pattern similar to that following the first wave, through May 2021, crossing above 1 in early April and peaking in mid-June; maximum R_cases_ in the third wave was 1.46 (CI: 1.38-1.53). R_cases_ dipped sharply from mid-June through late July, then climbed to a smaller peak in mid-to-late August. R_cases_ decreased through late August and September 2021, then remained approximately constant from late September through October 2021. Starting in early November 2021, R_cases_ increased rapidly throughout November 2021, reaching a maximum value of 3.33 (CI 2.83-3.97), then decreased rapidly through the end of December. R_cases_ increased slightly during January 2022, then remained constant during February and March. R_cases_ increased quickly in early to mid April 2022 (figure 1).

**Figure 1:**
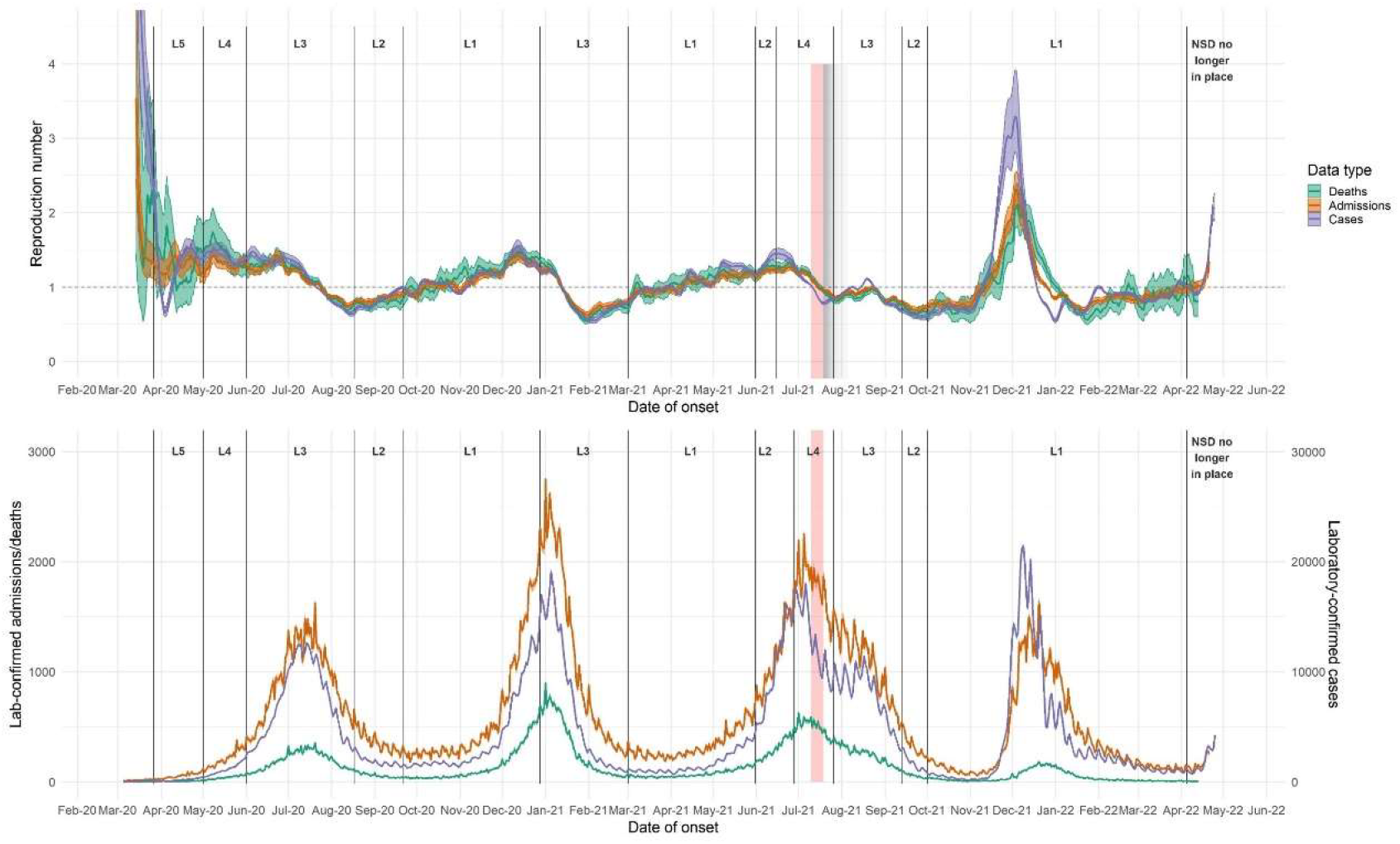
(upper panel) R estimates for each data endpoint, South Africa, based on (lower panel) national daily time series of rt-PCR-confirmed cases, hospitalisations, and deaths. R estimated using 7-day sliding windows, from early March 2020 through through 25 April. Results reflect median values (between imputations) of median R estimates and associated 2.5% and 97.5% credible intervals. L = Level. Red shaded areas indicate the period during which civil unrest caused severe disruptions to surveillance in KwaZulu-Natal and Gauteng provinces; grey shaded areas indicate gradually diminishing effects on R estimates.

**Table 1:** National average R for each consecutive lockdown level, with 2.5% and 97.5% credible intervals. Dates indicate the start of each period.

| Lockdown Level | R<sub>cases</sub> | R<sub>admissions</sub> | R<sub>deaths</sub> |
| --- | --- | --- | --- |
| Pre-lockdown | 2.32 (2.00,2.74) | 1.56 (1.38,1.78) | 1.88 (1.29,2.59) |
| L5 (27 March 2020) | 1.29 (1.24,1.34) | 1.27 (1.20,1.34) | 1.32 (1.14,1.50) |
| L4 (1 May 2020) | 1.40 (1.34,1.46) | 1.29 (1.24,1.35) | 1.35 (1.25,1.45) |
| L3 (1 June 2020) | 1.02 (1.02,1.03) | 1.03 (1.02,1.04) | 1.03 (1.01,1.05) |
| L2 (18 August 2020) | 0.83 (0.81,0.85) | 0.85 (0.83,0.87) | 0.79 (0.75,0.83) |
| L1 (21 September 2020) | 1.23 (1.19,1.26) | 1.21 (1.18,1.24) | 1.30 (1.25,1.35) |
| L3 (29 December 2020) | 0.85 (0.84,0.87) | 0.88 (0.86,0.89) | 0.86 (0.85,0.88) |
| L1 (1 March 2021) | 1.13 (1.12,1.15) | 1.07 (1.06,1.09) | 1.10 (1.07,1.13) |
| L2 (31 May 2021) | 1.42 (1.36,1.48) | 1.25 (1.21,1.29) | 1.28 (1.23,1.34) |
| L4 (16 June 2021) | 1.01 (1.00,1.02) | 1.05 (1.04,1.07) | 1.05 (1.04,1.07) |
| L3 (26 July 2021) | 0.91 (0.90,0.92) | 0.90 (0.88,0.91) | 0.87 (0.85,0.89) |
| L2 (13 September 2021) | 0.63 (0.60,0.67) | 0.72 (0.68,0.75) | 0.65 (0.60,0.70) |
| L1 (1 October 2021) | 1.00 (1.00,1.00) | 0.99 (0.98,1.00) | 0.97 (0.95,0.99) |
| Post-NSOD (5 April 2022) | 1.60 (1.51,1.68) | 1.26 (1.21,1.31) | 0.85 (0.66,1.08) |

Trends in R_cases_ between provinces were generally similar, though several provinces (Northern Cape, North West, and Free State) exhibited extended first waves. Estimates for Western Cape, Gauteng, and to a lesser extent KwaZulu-Natal indicate transitory increases in transmission during October 2020, prior to the onset of the second wave (figure 2 and supplementary materials section 1).

**Figure 2:**
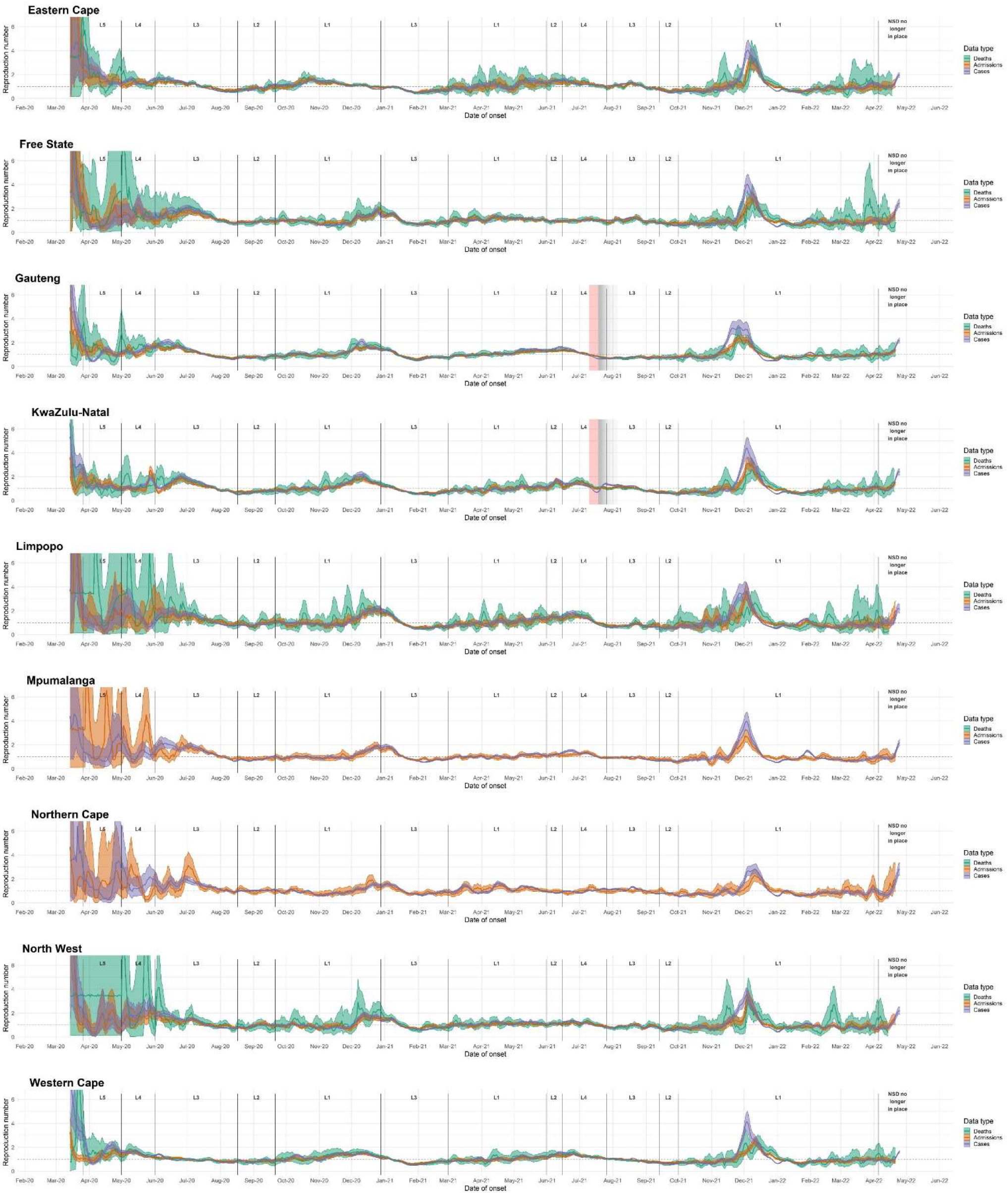
Province-level R estimates for each data endpoint from early March 2020 through 25 October 2022. R estimated on 7-day sliding windows. Results reflect median values (between imputations) of median R estimates and associated 2.5% and 97.5% credible intervals. L = Level. Red shaded areas indicate the period during which civil unrest caused severe disruptions to surveillance in KwaZulu-Natal and Gauteng provinces; grey shaded areas indicate gradually diminishing effects on R estimates.

While the timing of transmission trends varied substantially between provinces during the first wave, peak R_cases_ values during the second (Beta dominated) wave occurred at similar times in most provinces, except in the Eastern Cape, where the second wave started approximately four weeks earlier than in other provinces; in addition, Limpopo, Mpumalanga, Northern Cape, and Free State experienced peak transmission slightly later than the more-densely populated provinces of Western Cape, Gauteng, and KwaZulu-Natal.

The timing and shape of the third (Delta dominated) wave was more varied between provinces than the first two waves (figure 2 and supplementary materials section 1). Limpopo, Mpumalanga, North West, and Gauteng provinces experienced peak R_cases_ in late June or early July, while the Eastern Cape, KwaZulu-Natal, Western Cape, and Free State experienced peak R_cases_ in late July or August.

### Comparing R estimates based on different data endpoints

Trends in R estimates based on cases (R_cases_), hospitalizations (R_admissions_), and in-hospital deaths (R_deaths_) were generally similar during the first three waves, but R_cases_ diverged from R_admissions_ and R_deaths_ during the fourth wave, with the peak R_cases_ being higher than that from deaths and admissions. Throughout the epidemic, estimates based on deaths (and to a lesser extent admissions) were generally less stable and had wider credible intervals than estimates based on cases (see supplementary materials sections 2-3). Ratios between the three endpoints varied considerably over the course of the epidemic (see supplementary materials section 6).

Shortly after the peak of the first wave, in July 2020, R_cases_ dropped below R_admissions_ and R_deaths_; this occurred in several provinces as well as nationally. During the first wave, in mid-May 2020, R_deaths_ was higher than R_cases_, which was in turn higher than R_admissions_. In November 2020, when R_cases_ and R_admissions_ dipped, as well as in late December 2020 and early January 2021, R_deaths_ was higher than R_cases_ and R_admissions_. In six out of nine provinces, R_cases_ exceeded R_admissions_ and R_deaths_ during parts of June and July 2021.

In Gauteng, R_admissions_ had lower maxima in both waves than R_cases_ or R_deaths_; R_cases_ had similar maxima to R_deaths_ but maintained these values for shorter periods. R_cases_ reached lower post-wave minima than R_admissions_ or R_deaths_. In KwaZulu-Natal, R_admissions_ changed less drastically during the first two waves, with lower maxima and higher minima, than R_cases_ or R_deaths_. Civil unrest in Gauteng and KwaZulu-Natal during July 2021 caused substantial disruptions to laboratory and healthcare services, with corresponding dips in R estimates followed by compensatory increases. All three endpoints were affected, though R_cases_ was most affected.

During the fourth wave in late 2021 and early 2022, R_cases_ rose faster and higher than R_admissions_ or R_deaths_. R estimates based on the three data endpoints peaked within two days of one another in early December 2021, after which R_cases_ dropped more rapidly than R_admissions_, which in turn dropped faster than R_deaths_. Estimates based on the three endpoints converged again in mid-February 2022.

### Public versus private sector

Transmission patterns between clients of the public and private sectors were generally similar. Public and private sector R estimates differed most during the initial level five and four lockdowns, and in R_cases_ leading up to the peak of the fourth wave. At the end of the third wave in June/July 2021, and to a lesser extent at the end of the second wave, private-sector R dropped more rapidly than public-sector R, so that private-sector R was lower than public sector R (figure 3). This difference appears in estimates using all three data endpoints for the national-level analysis, as well as in Gauteng, Free State, and KwaZulu-Natal, and appeared in estimates based on cases in all provinces except the Western Cape (see supplementary materials section 3). During the fourth wave, R_cases_ in the private sector rose above R_cases_ in the public sector.

**Figure 3:**
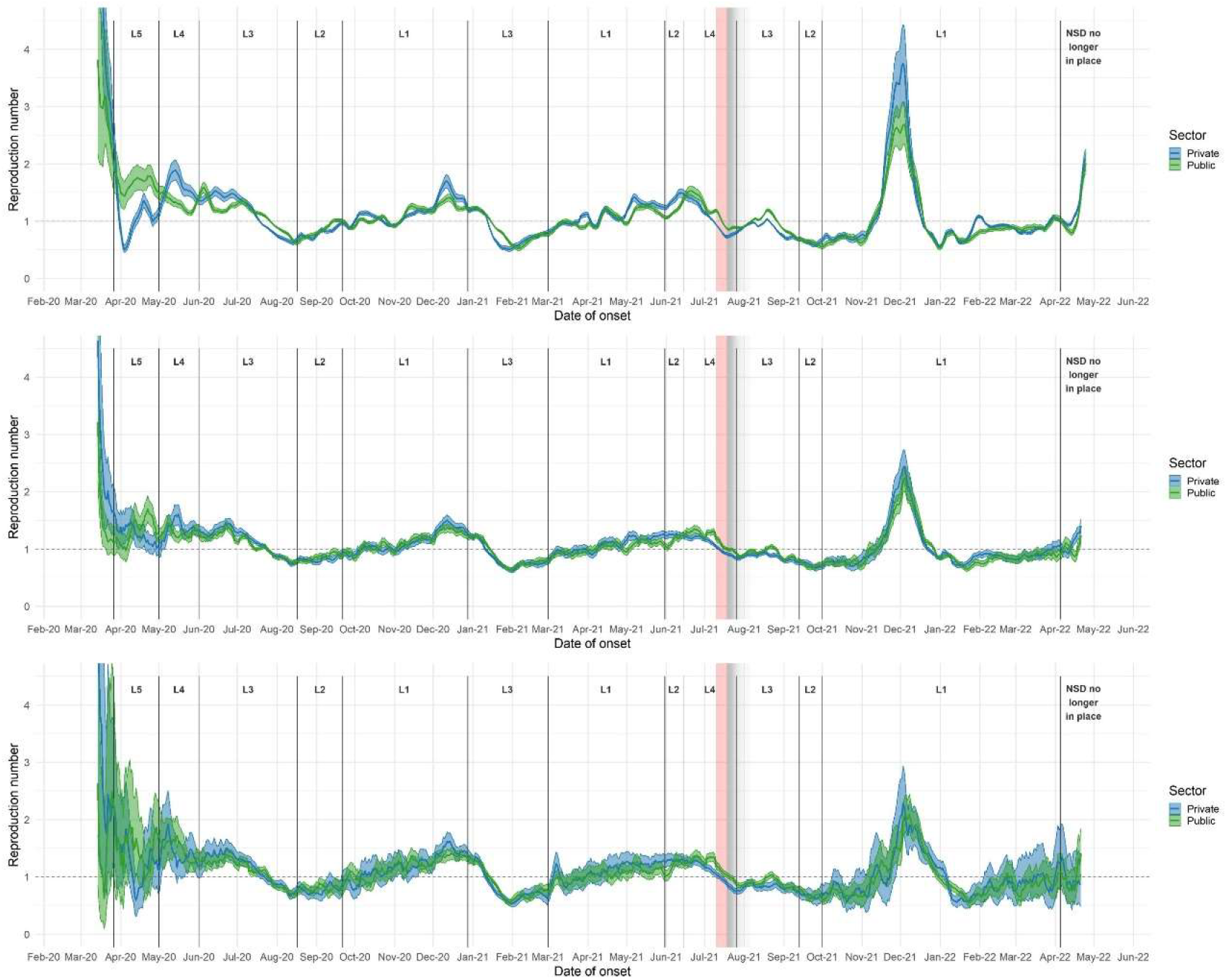
R estimates by sector, based on rt-PCR-confirmed COVID-19 cases (upper panel), hospitalizations (middle panel), and deaths (lower panel), South Africa. R estimates were generated using 7-day sliding windows. Results reflect median values (between imputations) of median R estimates and associated 2.5% and 97.5% credible intervals. L = Level. Red shaded areas indicate the period during which civil unrest caused severe disruptions to surveillance in KwaZulu-Natal and Gauteng provinces; grey shaded areas indicate gradually diminishing effects on R estimates.

## Discussion

### Overview

The initial level five lockdown in early 2020 had a substantial impact on transmission but was insufficient to bring R below one (table 1). It is difficult to disentangle the effects of subsequent lockdowns from those of increasing population immunity and other drivers of behavioral change. Even given high levels of preceding population immunity [31,32], the estimated peak R for the Omicron wave was substantially higher than previous waves. Average R values were lower in lower lockdown levels, probably because lockdown levels were increased in response to increasing transmission and lowered when transmission was deemed to be under control.

R estimates based on the three data endpoints were similar overall, although in practice hospitalisations and deaths provided slightly less timeous estimates. During our regular public reporting of R estimates [14], data updates on hospitalized cases and deaths were typically delayed by one to two weeks. Combined with longer truncation of hospitalized cases and deaths to account for late arriving data, this meant that estimates of R_cases_ were two to three weeks ahead of those for R_admissions_, and three weeks ahead of those for R_deaths_. R followed generally similar patterns by province, with notable exceptions including extended waves in some provinces, an early second wave in the Eastern Cape, and unrest-related changes to R estimates in Gauteng and KwaZulu-Natal during July and August 2021. R estimates in the private and public sectors were similar prior to the fourth wave, except during the level five and four lockdowns of early 2020, but diverged during the fourth wave in late 2021 and early 2022.

Our national-level R estimates are consistent with estimates from two other South African studies utilizing comparable methods [18,33]. McCarthy et. al. estimated R for the period before March 18^th^ 2020 (when the first movement restrictions were enacted) at 4.15 (CI: 3.60-4.74), using a generation interval estimate of 5.7 +-2.7 days, and ignoring importation status [33]; we estimated an R of 3.80 (CI: 3.03-4.92) for the same period. Roussouw obtained R trajectories consistent with ours, with R crossing 1 at approximately the same time as using our estimates, and peaking at similar values [18].

Provinces with drawn-out first waves – Northern Cape, North West, and Free State – are the three provinces with the lowest population density and smallest populations, possibly reflecting discontinuous epidemics in more isolated populations [34].

Events in some provinces, such as the civil unrest in KwaZulu-Natal and Gauteng provinces during July 2021, were associated with sudden drops in R estimates, followed by compensatory increases (see figures 1-3). In general, short-term decreases in incidence observation processes (such as disruptions to testing facilities or decreases in healthcare-seeking behavior) led to transitory dips in R estimates, followed by compensatory increases. This sort of dip and increase typically occurs for public holidays – e.g., in most provinces, Easter weekend 2021 coincided with a slight dip in R_admissions_ and R_cases_ (figure 2). In KwaZulu-Natal, R estimates based on admissions (R_admissions_) spiked following a nosocomial outbreak at a private hospital in early April [35] (figure 2). In addition, on 26 May 2020 a public hospital in Durban, KwaZulu-Natal, reported a spike of ∼100 admissions – this led to a spike, and subsequent dip, in R estimates for KZN. All of these factors highlight the fact that case-based estimates can be affected by changes in testing practice which could bias R estimates, particularly over the short term.

### Different data endpoints

R estimation using hospital admissions and deaths may prove valuable in scenarios where laboratory testing is limited or unavailable to the general public or if there is changing access to testing throughout the pandemic – e.g., policies restricting testing to severe cases during epidemic peaks such as those implemented in the Western Cape [36]. Numbers of hospitalisations should be more robust to changes in test availability, although changing hospitalization admission criteria may coincide with changes in the laboratory testing data; similarly, in-hospital deaths may change relative to incidence of infections due to improvements in treatment practices leading to reduced mortality over time. Overall, the general agreement of R estimates between endpoints prior to the fourth wave is encouraging, and points to the consistency of the imputation procedure. While agreement between endpoints would also suggest similarities in the levels of representativeness of each data endpoint, the ratios of incidence from the three time-series varied substantially over the time period considered (see supplementary materials section 6).

Stricter hospital admission practices near times of peak transmission, and corresponding relaxations in hospital admission practices following wave peaks, could have resulted in decreasing representativeness of hospitalizations (as a measure of incidence) during wave peaks and increasing representativeness following waves; this could explain the less extreme values and slower changes in R_admissions_ relative to R_cases_. On the other hand, testing seeking behavior may shift during waves, with heightened levels of concern during the beginning of waves leading to increases in test seeking and case confirmations. As wave peaks pass people may be less concerned, leading to a reduction in test seeking behavior and corresponding decrease in R estimates based on confirmed cases. Divergence between R_cases_ and R_admissions_ / R_deaths_ during the fourth wave was likely caused by a combination of the above factors with reduced severity of outcomes for people infected with the Omicron variant relative to the previously dominant Delta variant, causing the proportion of underlying infections which resulted in hospital admissions to decrease [37,38].

Were surveillance resources to be scaled back in future and the reliability of one or two of the data endpoints called into question, our results suggest that the remaining data endpoints could still be used to monitor changes in transmission, although robust surveillance of all three data endpoints should be maintained in some areas for validation purposes, particularly as increasing decoupling of COVID-19 cases, hospitalisations, and deaths is likely going forward.

### Public versus private sector

The similarities between public- and private-sector R estimates are of particular interest in light of several seroprevalence studies which suggest that clients of the public sector experienced higher levels of SARS-COV-2 transmission [7,22,39], as well as the under-representation of clients of the public sector in all three data endpoints relative to clients of the private sector – clients of the private sector make up approximately 17% of the South African population, while only 52% of recorded cases were in the private sector.

The most straightforward reason for the similarity of public and private sector R estimates is the fact that clients of the public and private sectors do not form two separate and isolated populations with regard to respiratory virus transmission – though there may have been less mixing during the initial highly-restrictive lockdown levels five and four, leading to differing R estimates during this period. In addition, several biases could have made it more difficult to detect differences in transmission: for example, healthcare-seeking behavior driven by heightened concern during periods of high incidence may have increased the proportion of infections which appeared in private sector data, inflating private-sector R estimates near peaks in R. This explanation is consistent with the higher R_cases_ based on private sector data during the fourth wave. Capacity limitations of public-sector healthcare providers and laboratory services (particularly during the first wave - see supplementary materials section 7 - when public-sector testing experienced substantial processing delays and backlogs) may also have led to decreasing representativeness of public-sector data near peaks in case and hospital admission numbers.

Unbiased R estimation requires that measures of disease incidence represent constant proportions of the true incidence of infections. In addition, the generation interval may have shifted over time with the appearance of new variants and changes in disease-related behaviors. Changes in laboratory capacity, including availability of test kits and reagents, levels of population immunity (whether pre-existing, infection-induced, or vaccine-induced), healthcare-seeking behavior, treatment of COVID-19 disease, circulating SARS-COV-2 strains, and data collection practices may all bias R estimates.

Furthermore, a number of factors may have altered mortality outcomes over time, including the introduction of dexamethasone treatment in mid-June 2020, the use of oxygen administration via high flow nasal cannula, changes in quality of healthcare provided if health systems are overwhelmed, changing levels of vaccine- and infection-induced immunity, and potential differences in severity between initially circulating viruses and the different variants which dominated the second, third, and fourth waves. Combined, these factors may lead to perturbations in the time series data that are unrelated to transmission. In addition, the distributions of delays between symptom onset and case report/admission/death may change over time, which would affect the accuracy of adjustments for right-censoring at the end of the time series. Due to the impact of civil unrest, which led to reductions in testing rates in KwaZulu-Natal and Gauteng provinces in July 2021, trends in R estimates during that time for these provinces, as well as nationally, should be interpreted with caution.

The agreement of R estimates based on the three data endpoints prior to the fourth wave, along with the fact that many of the likely biases would affect only one or two of the three endpoints, suggests that either the above biases were relatively small and/or together moved the three data endpoints in similar ways.

Early (pre-lockdown) incidence included a substantial portion of imported cases and our early R estimates are likely not an accurate reflection of transmission trends; the sharp drop in R estimates following the initial border closures may be attributed in part to the sudden decrease in imported cases (see McCarthy *et al*. [33] for early R estimates incorporating data on importation status).

Future work could include comparison with other R estimation methods [30,40], methods to correct for holidays and disruptions to surveillance processes (e.g. due to civil unrest), and disentangling the role of immunity from trends in R.

## Conclusion

We conducted a robust process of R estimation for COVID-19 in South Africa. The use of high-resolution data allowed us to directly compare estimates based on different data endpoints, since all estimates were based on symptom onset dates, to compare estimates between the public and private sectors, and to exclude antigen testing due to issues with data quality and reporting completeness.

We found that different data endpoints yielded similar R estimates during the first three waves but diverged during the fourth wave, suggesting that – while useful R estimates could potentially be obtained from only one or two data endpoints, particularly if virus severity remained unchanged – decision makers should where possible consider R estimates using multiple data endpoints. We found similar R estimates using public and private sector data, although clients of the public sector were heavily underrepresented in the data, and private sector R_cases_ was higher than public sector R_cases_ during the fourth wave.

Although R estimates provide limited resolution for understanding the drivers of transmission, the estimates presented here served a valuable role in the South African COVID-19 response efforts by providing routine monitoring of transmission trends.

## Data Availability

Time series data used for R estimation, along with relevant code for reproducing results based on this data, are available upon reasonable request to the authors.

https://zenodo.org/deposit/6948468

https://github.com/SACEMA/R_est_sa

## Acknowledgements

We thank the numerous individuals and organizations involved in collecting and curating the DATCOV and NMC line list datasets (NMC epidemiology team: A. Moipone Shonhiwa, G. Ntshoe, J. Ebonwu, L. Motsuku, L. Shuping, M. Muchengeti, J. Kleynhans, G. Hunt, V. Odhiambo Olago, H. Ismail, N. Govender, A. Mathews, V. Essel, V. Msimang, T. Kufa-Chakezha, N. Villyen Motaze, N. Mayet, T. Mmaborwa Matjokotja, M. Neti, T. Arendse, T. Lamola, I. Matiea, D. Muganhiri, B. Ndlovu, K. Ravhuhali, E. Ramutshila, S. Mhlanga, A. Mzoneli, N. Naran, T. Whitbread, M. Moeti, C. Iwu, E. Mathatha, F. Gavhi, M. Makamu, M. Makhubele, S. Mdleleni, B. Chiger, and J. Kleynhans; information technology team: T. Mukange, T. Bell, L. Darwin, F. McKenna, N. Munava, M. Raza Bano, T. Ngobeni; DATCOV team: L. Blumberg, R. Kai, S. Dyasi, T. Arendse, M. Masha, B. Cowper, K. Skhosana, F. Malomane, M. Blom, A. Mzoneli, S. Mhlanga, B. Ali, C. Mudara, L. Ozougwu, R. Welch, N. Mfongeh, P. Manana, Y. Mangwane, M. Mokgosana, T. Buthelezi, P. Makwene, M. Dryden, C. Vika).

## References

1. Childs SJ. Quantification of the South African Lockdown Regimes, for the SARS-CoV-2 Pandemic, and the Levels of Immunity They Require to Work. medRxiv. 2020; 2020.07.11.20151555. doi:10.1101/2020.07.11.20151555

2. Direct and Indirect Health Effects of Lockdown in South Africa. In: Center For Global Development [Internet]. [cited 27 May 2021]. Available: https://www.cgdev.org/publication/direct-and-indirect-health-effects-lockdown-south-africa

3. Regulations and Guidelines - Coronavirus COVID-19 South African Government. [cited 20 Jul 2021]. Available: https://www.gov.za/covid-19/resources/regulations-and-guidelines-coronavirus-covid-19

4. Network for Genomic Surveillance in South Africa (NGS-SA). SARS-CoV-2 Sequencing Update. National Institute for Communicable Diseases, South Africa; 2021. Available: https://www.nicd.ac.za/wp-content/uploads/2022/01/Update-of-SA-sequencing-data-from-GISAID-30-Dec-2021_dash.pdf

5. Statistics South Africa. General Household Survey 2018. Statistics South Africa; 2019 May p. 203. Report No.: 318. Available: https://www.statssa.gov.za/publications/P0302/P03022019.pdf

6. Söderlund N, Hansl B. Health insurance in South Africa: an empirical analysis of trends in risk-pooling and ef?ciency following deregulation. Health Policy Plan. 2000;15: 378–385. doi:10.1093/heapol/15.4.378

7. Shaw JA, Meiring M, Cummins T, Chegou NN, Claassen C, Plessis ND, et al. Higher SARS-CoV-2 seroprevalence in workers with lower socioeconomic status in Cape Town, South Africa. PLOS ONE. 2021;16: e0247852. doi:10.1371/journal.pone.0247852

8. Chatterjee A, UNU-WIDER. Measuring wealth inequality in South Africa: An agenda. 45th ed. UNU-WIDER; 2019. doi:10.35188/UNU-WIDER/2019/679-1

9. Maphumulo WT, Bhengu BR. Challenges of quality improvement in the healthcare of South Africa post-apartheid: A critical review. Curationis. 2019;42: 1901. doi:10.4102/curationis.v42i1.1901

10. Hawkins RB, Charles EJ, Mehaffey JH. Socio-economic status and COVID-19–related cases and fatalities. Public Health. 2020;189: 129–134. doi:10.1016/j.puhe.2020.09.016

11. JMIR Public Health and Surveillance - Novel Coronavirus in Cape Town Informal Settlements: Feasibility of Using Informal Dwelling Outlines to Identify High Risk Areas for COVID-19 Transmission From A Social Distancing Perspective. [cited 24 Jun 2021]. Available: https://publichealth.jmir.org/2020/2/e18844/

12. Cori A, Ferguson NM, Fraser C, Cauchemez S. A New Framework and Software to Estimate Time-Varying Reproduction Numbers During Epidemics. American Journal of Epidemiology. 2013;178: 1505–1512. doi:10.1093/aje/kwt133

13. Fraser C. Estimating Individual and Household Reproduction Numbers in an Emerging Epidemic. PLOS ONE. 2007;2: e758. doi:10.1371/journal.pone.0000758

14. NICD. THE DAILY COVID-19 EFFECTIVE REPRODUCTIVE NUMBER (R) IN SOUTH AFRICA. National Institute for Communicable Diseases; 2021 May p. 13. Available: https://www.nicd.ac.za/wp-content/uploads/2021/08/COVID-19-Effective-Reproductive-Number-in-South-Africa-week-32.pdf

15. Garba SM, Lubuma JM-S, Tsanou B. Modeling the transmission dynamics of the COVID-19 Pandemic in South Africa. Mathematical Biosciences. 2020;328: 108441. doi:10.1016/j.mbs.2020.108441

16. Giandhari J, Pillay S, Wilkinson E, Tegally H, Sinayskiy I, Schuld M, et al. Early transmission of SARS-CoV-2 in South Africa: An epidemiological and phylogenetic report. medRxiv. 2020 [cited 27 May 2021]. doi:10.1101/2020.05.29.20116376

17. Mbuvha R, Marwala T. Bayesian inference of COVID-19 spreading rates in South Africa. PLOS ONE. 2020;15: e0237126. doi:10.1371/journal.pone.0237126

18. Roussouw L. Estimating the Effective Reproduction Number of COVID-19 in South Africa. 2021. Available: https://unsupervised.online/static/covid-19/estimating_r_za.html

19. Olivier LE, Craig IK. An epidemiological model for the spread of COVID-19: A South African case study. arXiv e-prints. 2020;2005: arXiv:2005.08012.

20. Musa SS, Zhao S, Wang MH, Habib AG, Mustapha UT, He D. Estimation of exponential growth rate and basic reproduction number of the coronavirus disease 2019 (COVID-19) in Africa. Infectious Diseases of Poverty. 2020;9: 96. doi:10.1186/s40249-020-00718-y

21. May 26 AAP, Doi 2021. Covid-19: Estimates for South Africa. In: Covid-19 [Internet]. [cited 27 May 2021]. Available: https://epiforecasts.io/covid/posts/national/south-africa/

22. George JA, Khoza S, Mayne E, Dlamini S, Kone N, Jassat W, et al. Sentinel seroprevalence of SARS-CoV-2 in the Gauteng province, South Africa August to October 2020. Infectious Diseases (except HIV/AIDS); 2021 Apr. doi:10.1101/2021.04.27.21256099

23. Thompson RN, Stockwin JE, van Gaalen RD, Polonsky JA, Kamvar ZN, Demarsh PA, et al. Improved inference of time-varying reproduction numbers during infectious disease outbreaks. Epidemics. 2019;29: 100356. doi:10.1016/j.epidem.2019.100356

24. NICD. NMC COVID-19 DOCUMENTS. National Institute for Communicable Diseases, South Africa; 2020. Available: https://www.nicd.ac.za/nmc-overview/nmc-covid-19-documents/

25. Pulliam JRC, van Schalkwyk C, Govender N, von Gottberg A, Cohen C, Groome MJ, et al. Increased risk of SARS-CoV-2 reinfection associated with emergence of Omicron in South Africa. Science. 2022;376: eabn4947. doi:10.1126/science.abn4947

26. NICD National COVID-19 Hospital Surveillance. National Institute for Communicable Diseases; 2021 Jun p. 5. Available: https://www.nicd.ac.za/wp-content/uploads/2021/07/DATCOV-National-report-20210630.pdf

27. Cohen C, Kleynhans J, Gottberg A von, McMorrow ML, Wolter N, Bhiman JN, et al. SARS-CoV-2 incidence, transmission and reinfection in a rural and an urban setting: results of the PHIRST-C cohort study, South Africa, 2020-2021. 2021 Jul p. 2021.07.20.21260855. doi:10.1101/2021.07.20.21260855

28. Buuren S van, Groothuis-Oudshoorn K, Vink G, Schouten R, Robitzsch A, Rockenschaub P, et al. mice: Multivariate Imputation by Chained Equations. 2021. Available: https://CRAN.R-project.org/package=mice

29. Kleinke K. kkleinke/countimp. 2020. Available: https://github.com/kkleinke/countimp

30. Gostic KM, McGough L, Baskerville EB, Abbott S, Joshi K, Tedijanto C, et al. Practical considerations for measuring the effective reproductive number, Rt. PLOS Computational Biology. 2020;16: e1008409. doi:10.1371/journal.pcbi.1008409

31. Madhi SA, Kwatra G, Myers JE, Jassat W, Dhar N, Mukendi CK, et al. Population Immunity and Covid-19 Severity with Omicron Variant in South Africa. N Engl J Med. 2022;386: 1314–1326. doi:10.1056/NEJMoa2119658

32. Kleynhans J, Tempia S, Wolter N, von Gottberg A, Bhiman J, Buys A, et al. SARS-CoV-2 Seroprevalence in a Rural and Urban Household Cohort during First and Second Waves of Infections, South Africa, July 2020–March 2021. Emerging Infectious Disease journal. 2021;27. doi:10.3201/eid2712.211465

33. Mccarthy K, Tempia S, Kufa T, Kleynhans J, Wolter N, Jassat W, et al. The Importation and Establishment of Community Transmission of SARS-CoV-2 During the First Eight Weeks of the South African COVID-19 Epidemic. Rochester, NY: Social Science Research Network; 2021 Feb. Report No.: ID 3792114. doi:10.2139/ssrn.3792114

34. Statistics South Africa. Mid-year population estimates 2020. Statistics South Africa; 2020 Jul p. 35. Report No.: 302. Available: http://www.statssa.gov.za/publications/P0302/P03022020.pdf

35. Ministerial Advisory Committee (MAC) on COVID-19. St Augustine Hospital Outbreak of COVID-19 - Interim Report. National Department of Health, South Africa; 2020 May p. 2. Available: https://sacoronavirus.co.za/wp-content/uploads/2020/08/Memo_Advisory-St-Augustine-interim-report-final.pdf

36. Mahomed H, Gilson L, Boulle A, Davies M-A, Khan S, Carthy KM, et al. The evolution of the COVID-19 pandemic and health system responses in South Africa and the Western Cape Province – how decision-making was supported by data. District Health Barometer 2019/2020. Health Systems Trust; 2020. p. 18. Available: https://www.hst.org.za/publications/District%20Health%20Barometers/DHB%202019-20%20Section%20A,%20chapter%208%20-%20COVID-19%20pandemic.pdf

37. Wolter N, Jassat W, Walaza S, Welch R, Moultrie H, Groome M, et al. Early assessment of the clinical severity of the SARS-CoV-2 omicron variant in South Africa: a data linkage study. The Lancet. 2022;399: 437–446. doi:10.1016/S0140-6736(22)00017-4

38. Davies M-A, Kassanjee R, Rousseau P, Morden E, Johnson L, Solomon W, et al. Outcomes of laboratory-confirmed SARS-CoV-2 infection in the Omicron-driven fourth wave compared with previous waves in the Western Cape Province, South Africa. Tropical Medicine & International Health. 2022;27: 564–573. doi:10.1111/tmi.13752

39. Vermeulen M, Mhlanga L, Sykes W, Coleman C, Pietersen N, Cable R, et al. Prevalence of anti-SARS-CoV-2 antibodies among blood donors in South Africa during the period January-May 2021. 2021. doi:10.21203/rs.3.rs-690372/v1

40. Abbott S, Hellewell J, Sherratt K, Gostic K, Hickson J, Badr HS, et al. EpiNow2: Estimate Real-Time Case Counts and Time-Varying Epidemiological Parameters. 2020. Available: https://CRAN.R-project.org/package=EpiNow2

