## Supplementary materials for "Estimating the time-varying reproduction number for COVID-19 in South Africa during the first four waves using multiple measures of incidence for public and private sectors across four waves"

Contents:

1. Imputed time series
2. R estimates with different choices for width of sliding window
3. R estimates per province, per sector, with 21-day sliding window
4. Maximum and minimum R estimates per wave
5. Choice of parameters for imputation and R estimation procedures
6. Ratios between data endpoints
7. Tests conducted per sector

#### 1. Imputed time series by province

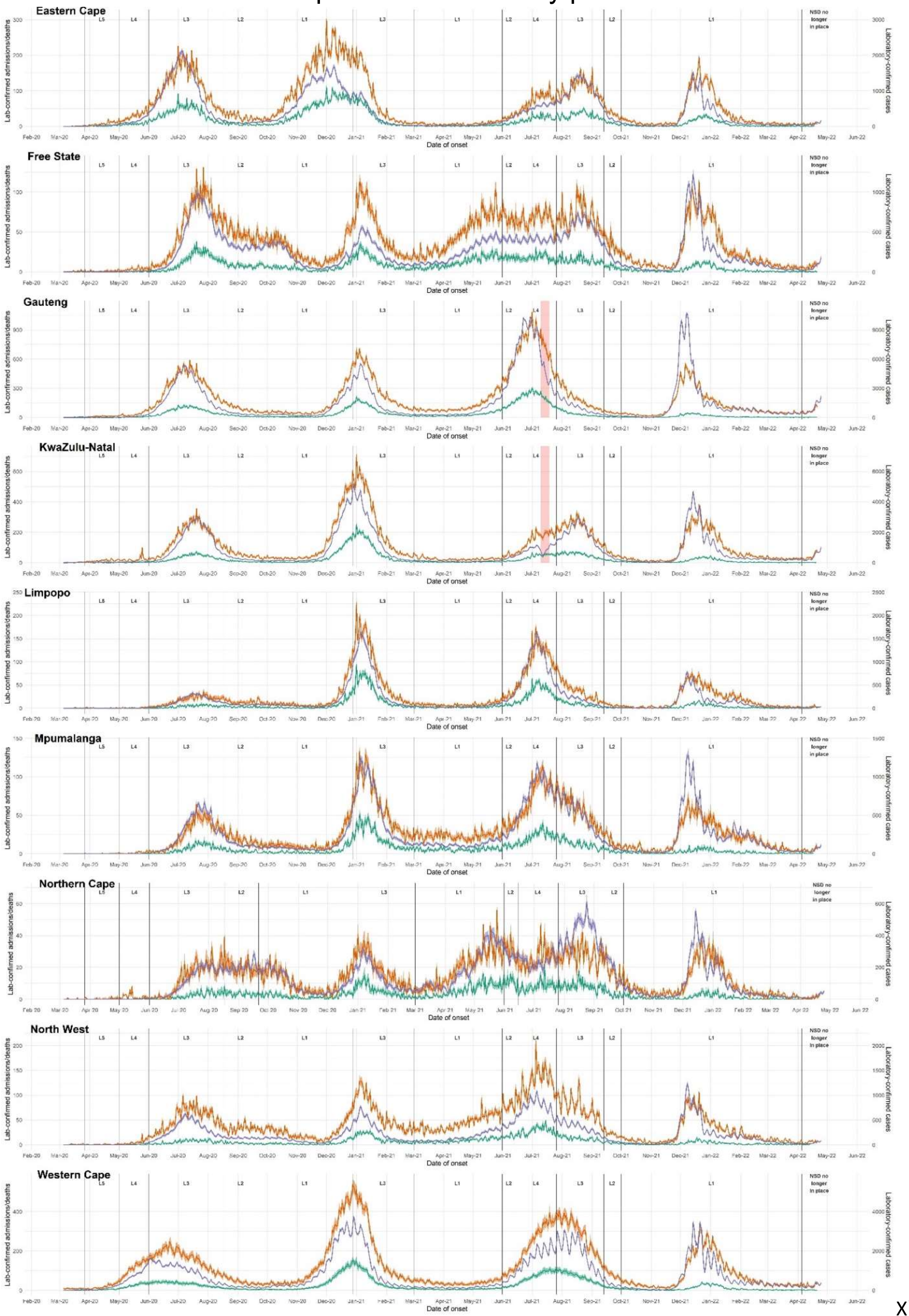


X`X

*Figure 1: Time series of laboratory-confirmed COVID-19 cases, hospitalizations, and deaths, by imputed dates of symptom onset, for each South African province, as used in our R estimation procedures. Plotted are median values, between imputations, with 2.5% and 97.5% quantiles.*

### 2. R estimates with different choices for width of sliding window

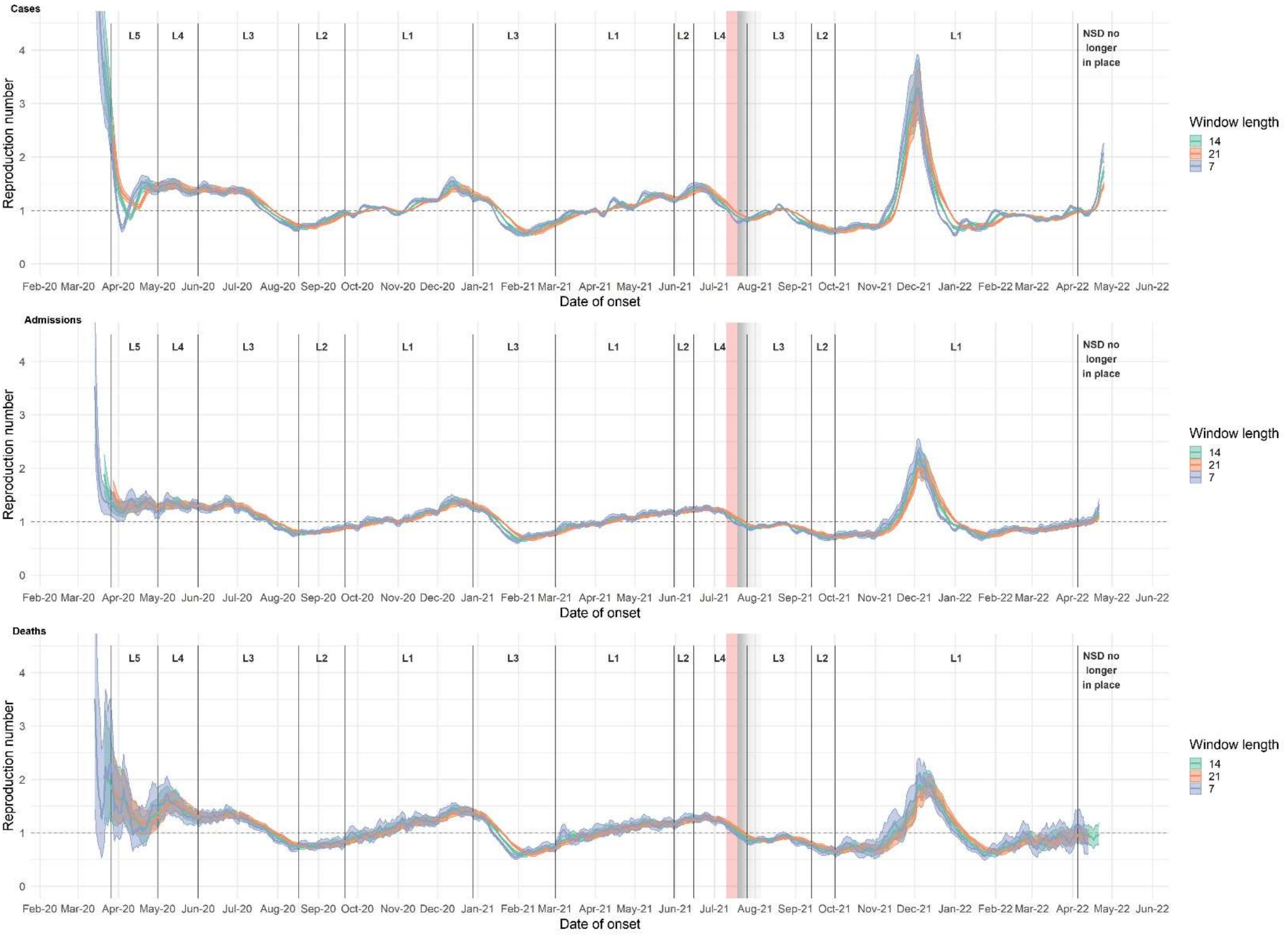


*Figure 2 R estimates based on rt-PCR-confirmed cases (upper panel), hospital admissions (middle panel) and hospital-associated deaths (lower panel) using 7-day, 14-day, and 21-day sliding windows. Results reflect median values (between imputations) of median R estimates and associated 2.5% and 97.5% credible intervals.*

#### 3. R estimates per province, per sector, with 21-day sliding window

##### a. National

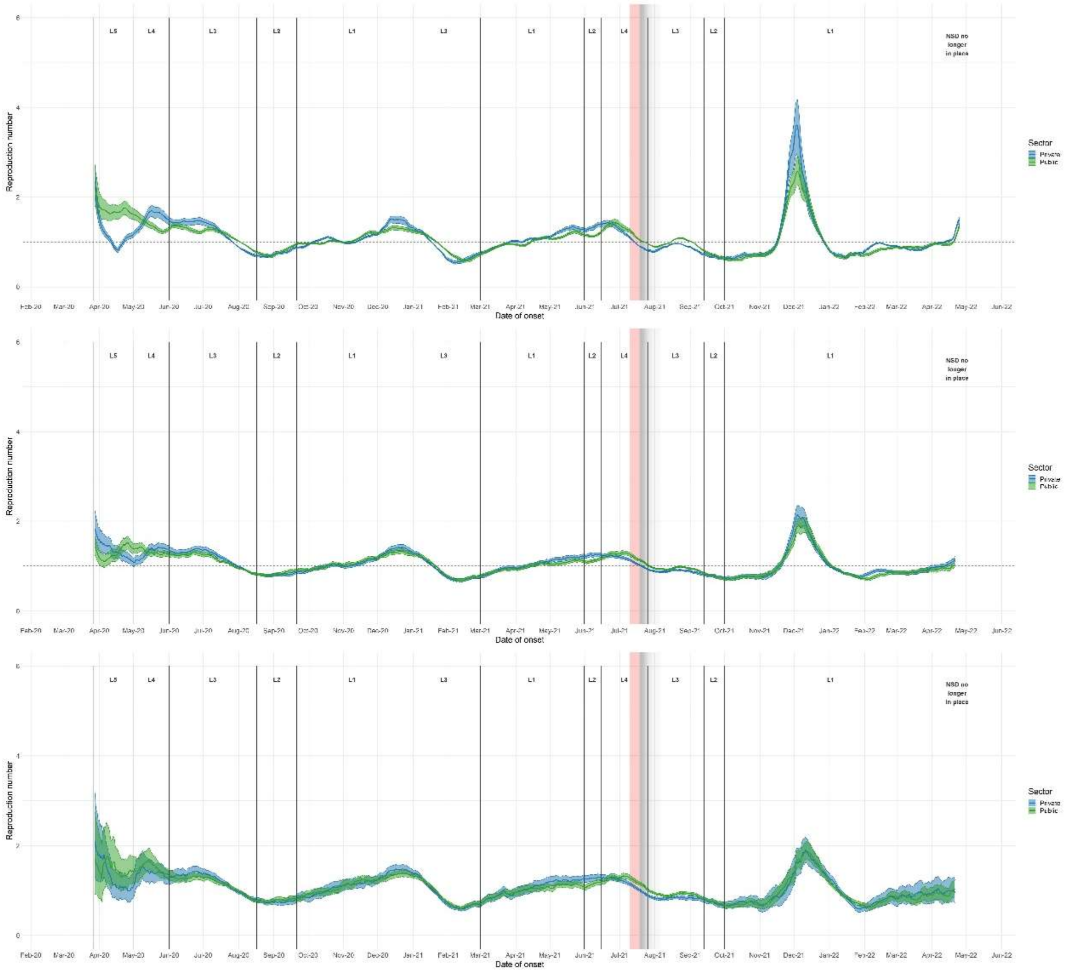


*Figure 3: R estimated separately for public and private sector data, based on cases (upper panel), admissions (middle panel) and deaths (lower panel), using 21-day sliding windows. Plotted are median values, between imputations, with 2.5% and 97.5% credible intervals.*

#### a. Eastern Cape

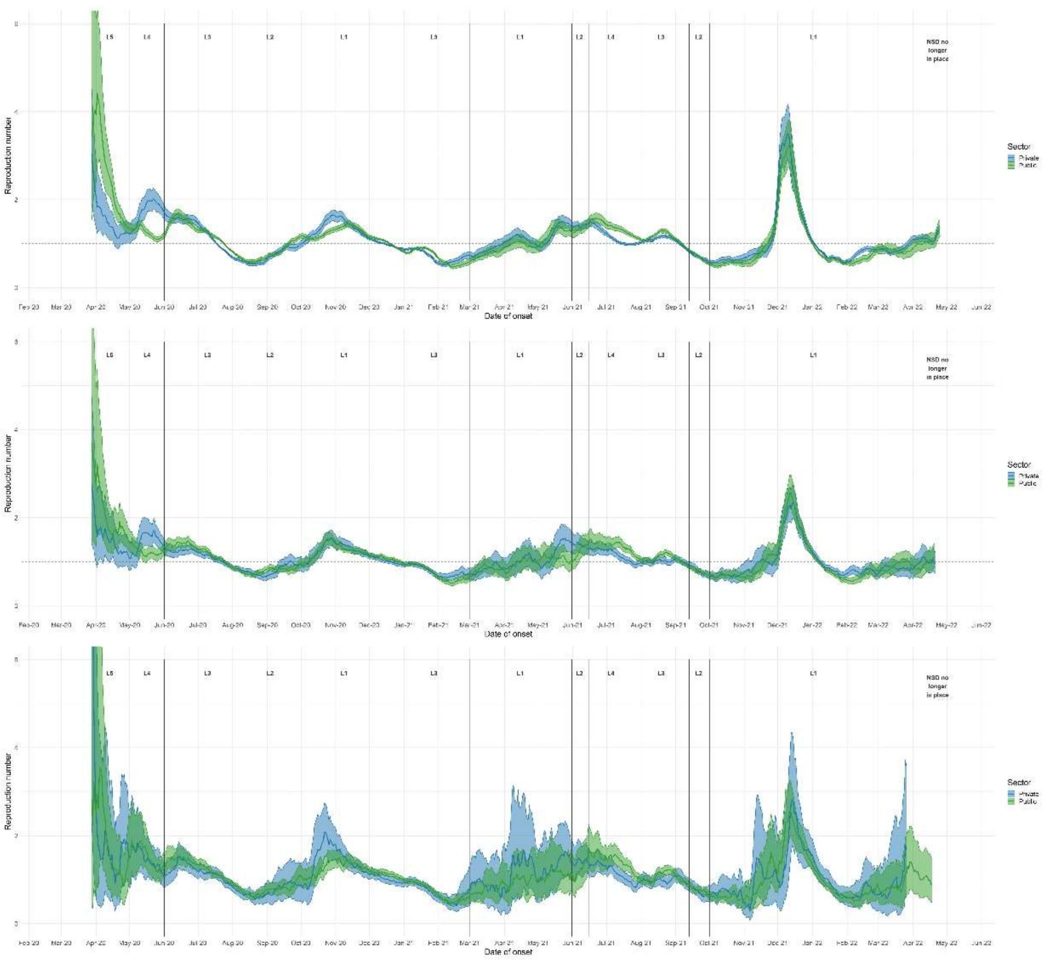


*Figure 4: R estimated separately for public and private sector data, based on cases (upper panel), admissions (middle panel) and deaths (lower panel), using 21-day sliding windows, Eastern Cape. Plotted are median values, between imputations, with 2.5% and 97.5% credible intervals.*

#### a. Free State

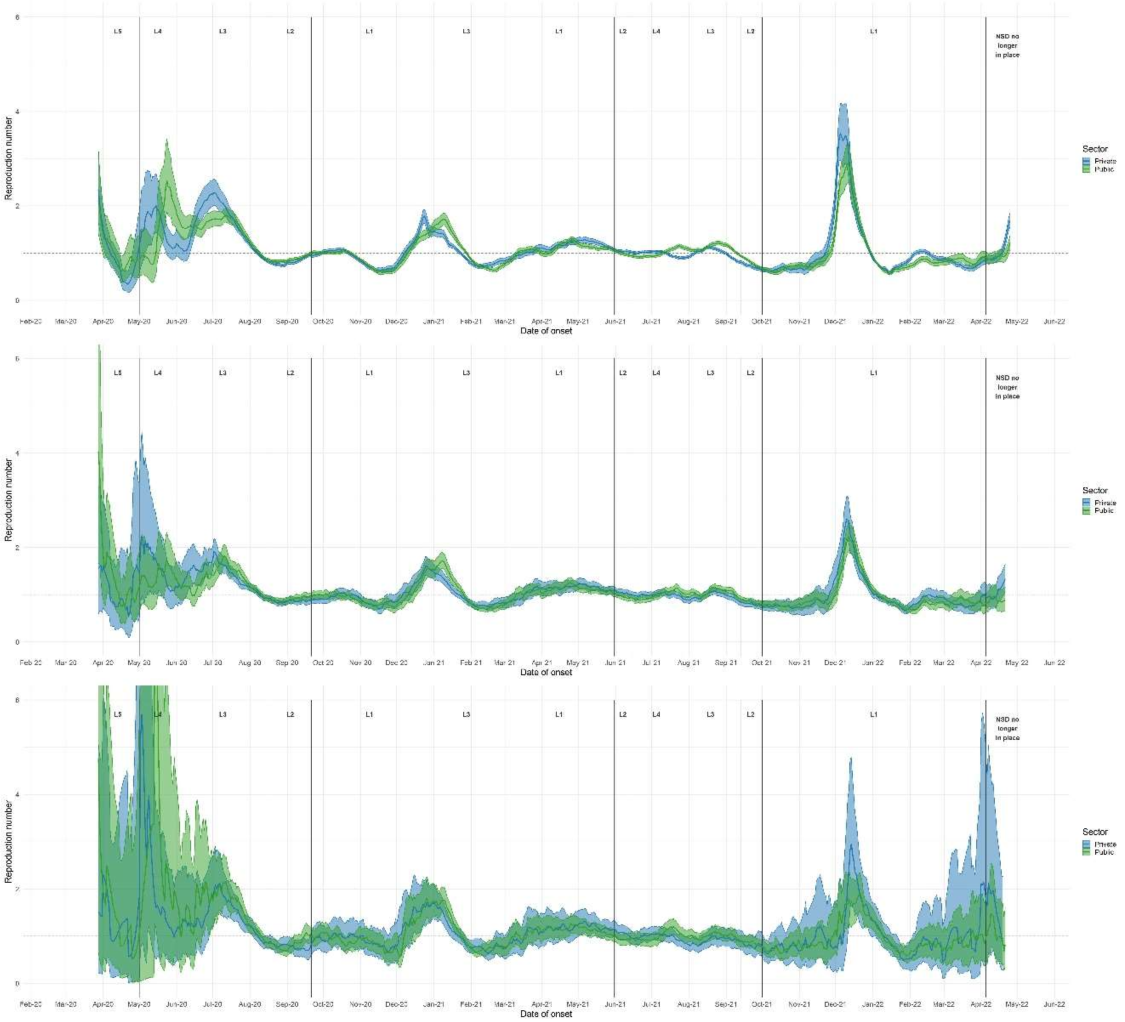


*Figure 5: R estimated separately for public and private sector data, based on cases (upper panel), admissions (middle panel) and deaths (lower panel), using 21-day sliding windows, Free State. Plotted are median values, between imputations, with 2.5% and 97.5% credible intervals.*

#### b. Gauteng

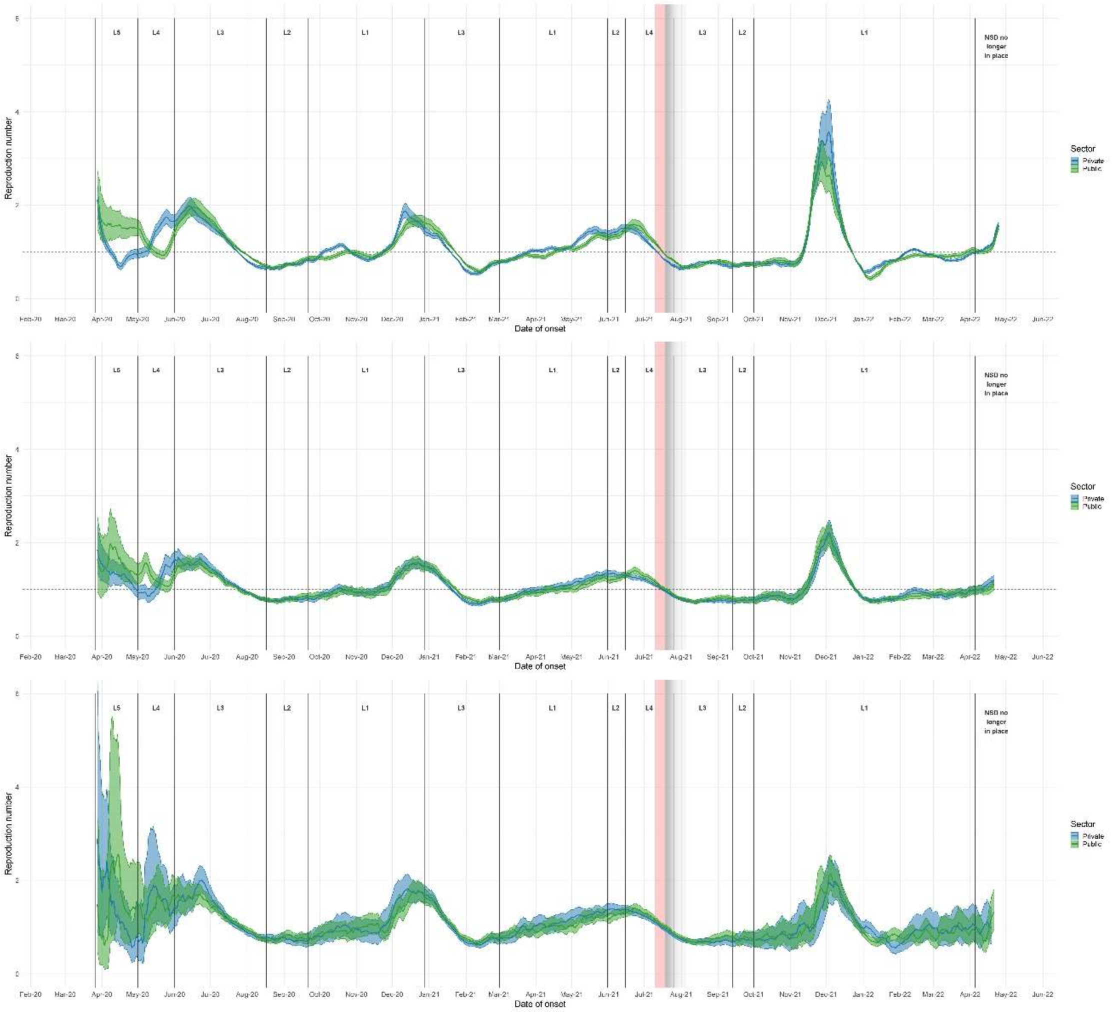


*Figure 6: R estimated separately for public and private sector data, based on cases (upper panel), admissions (middle panel) and deaths (lower panel), using 21-day sliding windows, Gauteng. Plotted are median values, between imputations, with 2.5% and 97.5% credible intervals.*

c. KwaZulu-Natal

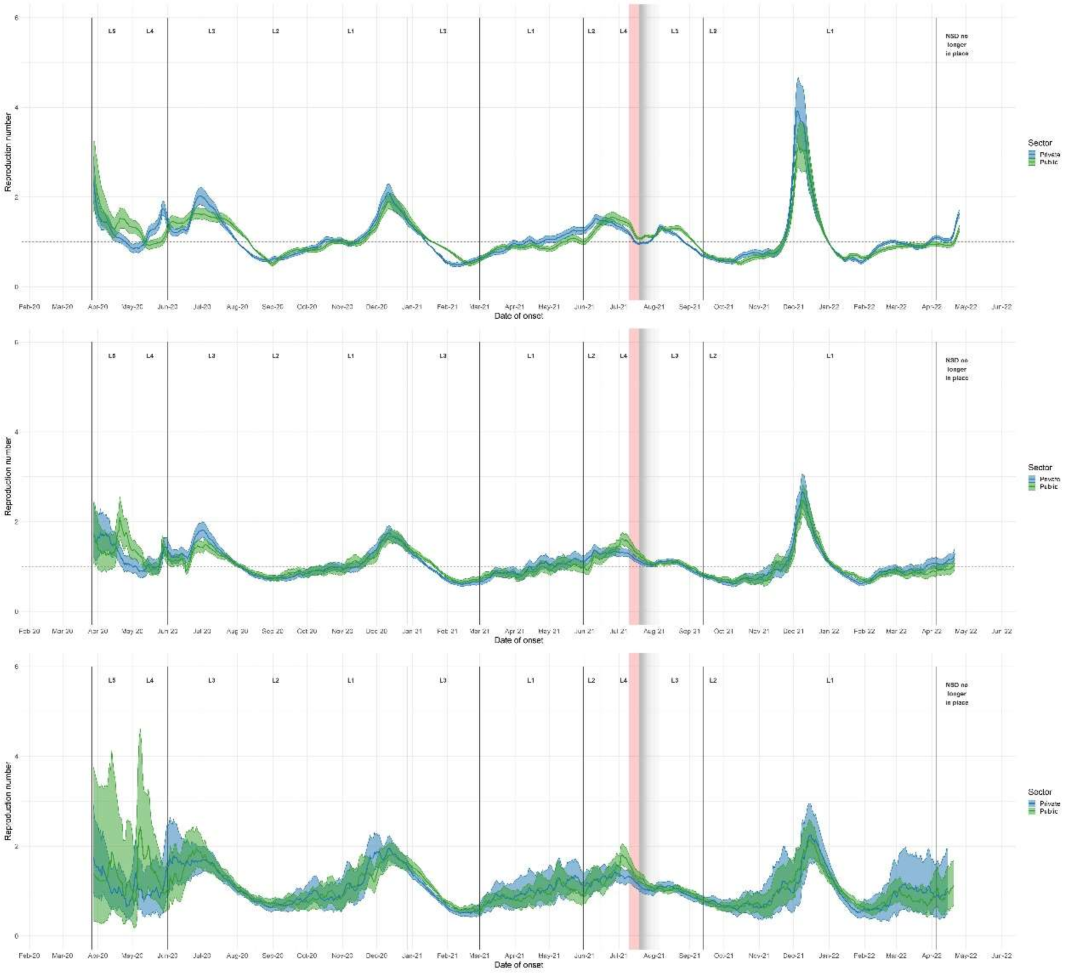


*Figure 7: R estimated separately for public and private sector data, based on cases (upper panel), admissions (middle panel) and deaths (lower panel), using 21-day sliding windows, KwaZulu-Natal. Plotted are median values, between imputations, with 2.5% and 97.5% credible intervals.*

#### d. Limpopo

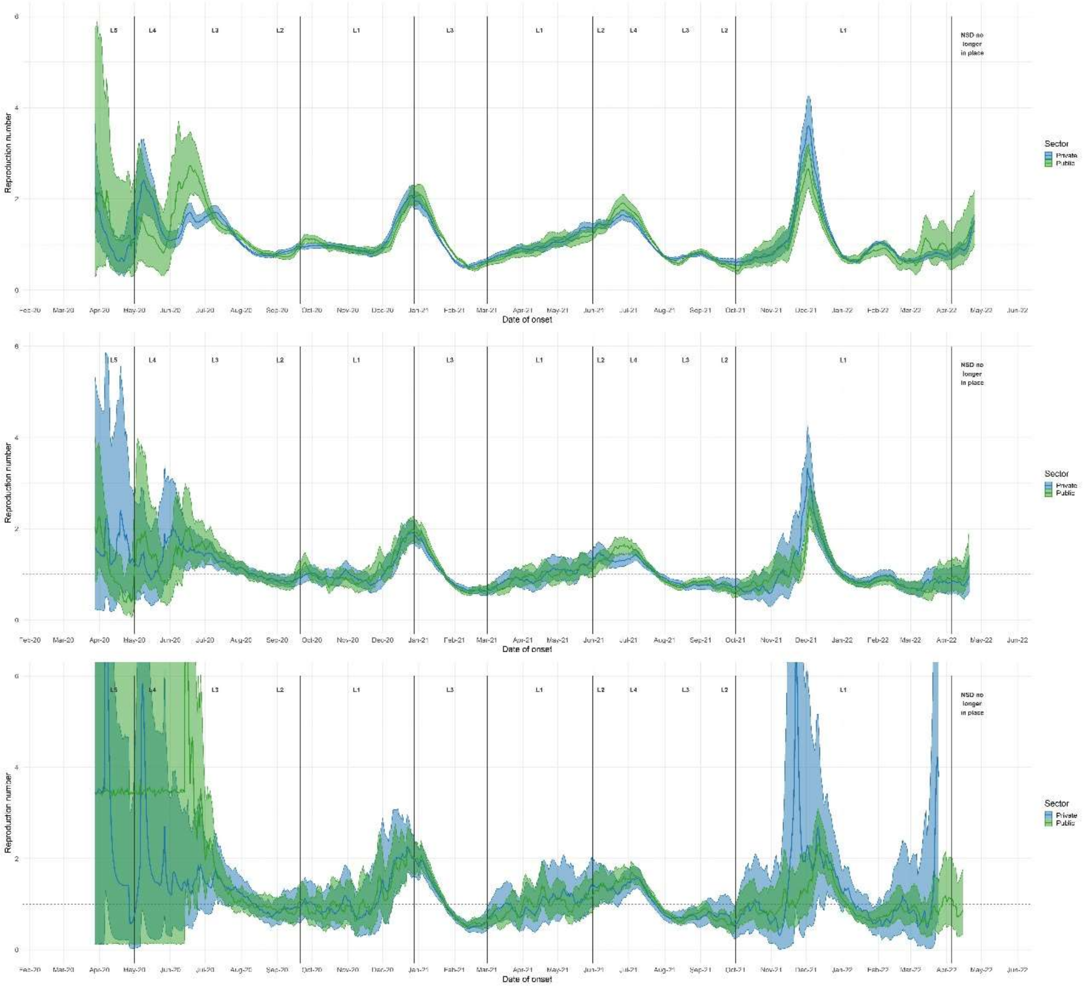


*Figure 8: R estimated separately for public and private sector data, based on cases (upper panel), admissions (middle panel) and deaths (lower panel), using 21-day sliding windows, Limpopo. Plotted are median values, between imputations, with 2.5% and 97.5% credible intervals.*

#### e. Mpumalanga

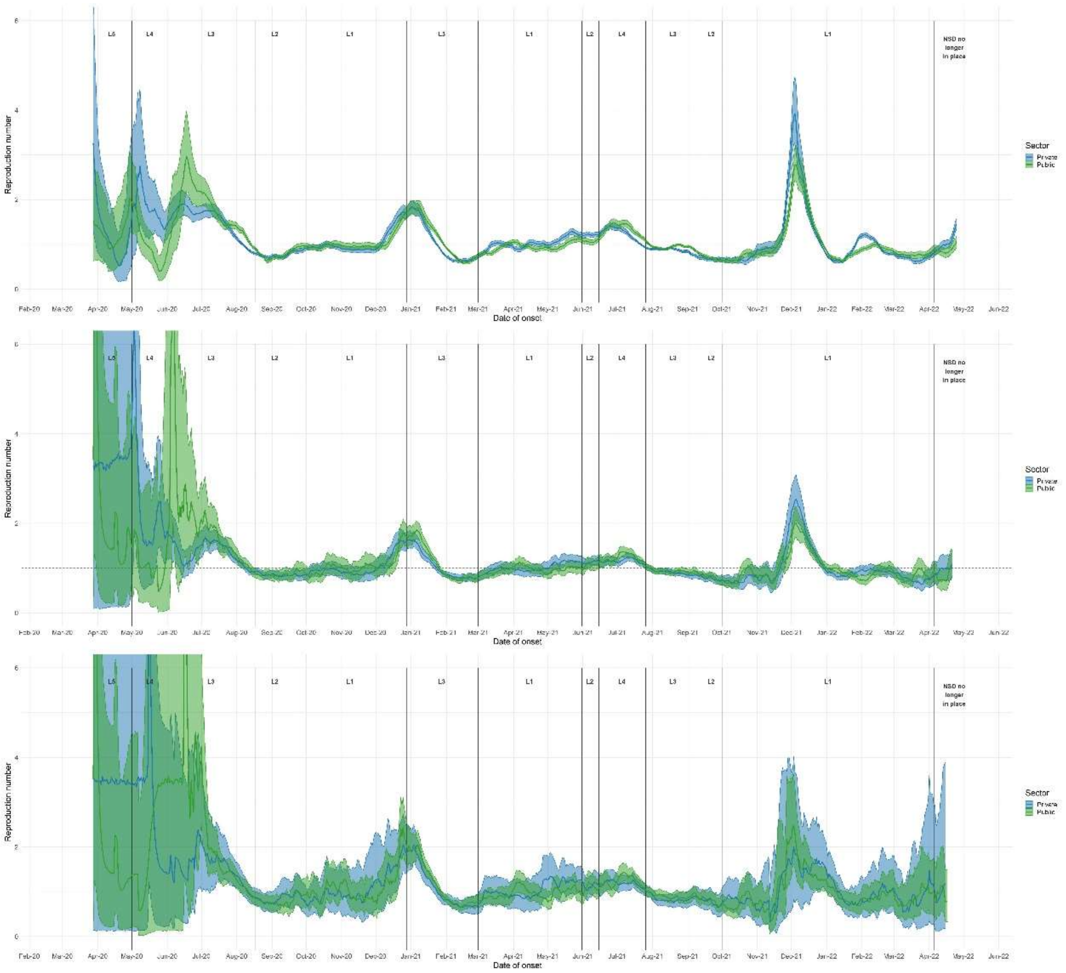

*Figure 9: R estimated separately for public and private sector data, based on cases (upper panel), admissions (middle panel) and deaths (lower panel), using 21-day sliding windows, Mpumalanga. Plotted are median values, between imputations, with 2.5% and 97.5% credible intervals.*

#### f. Northern Cape

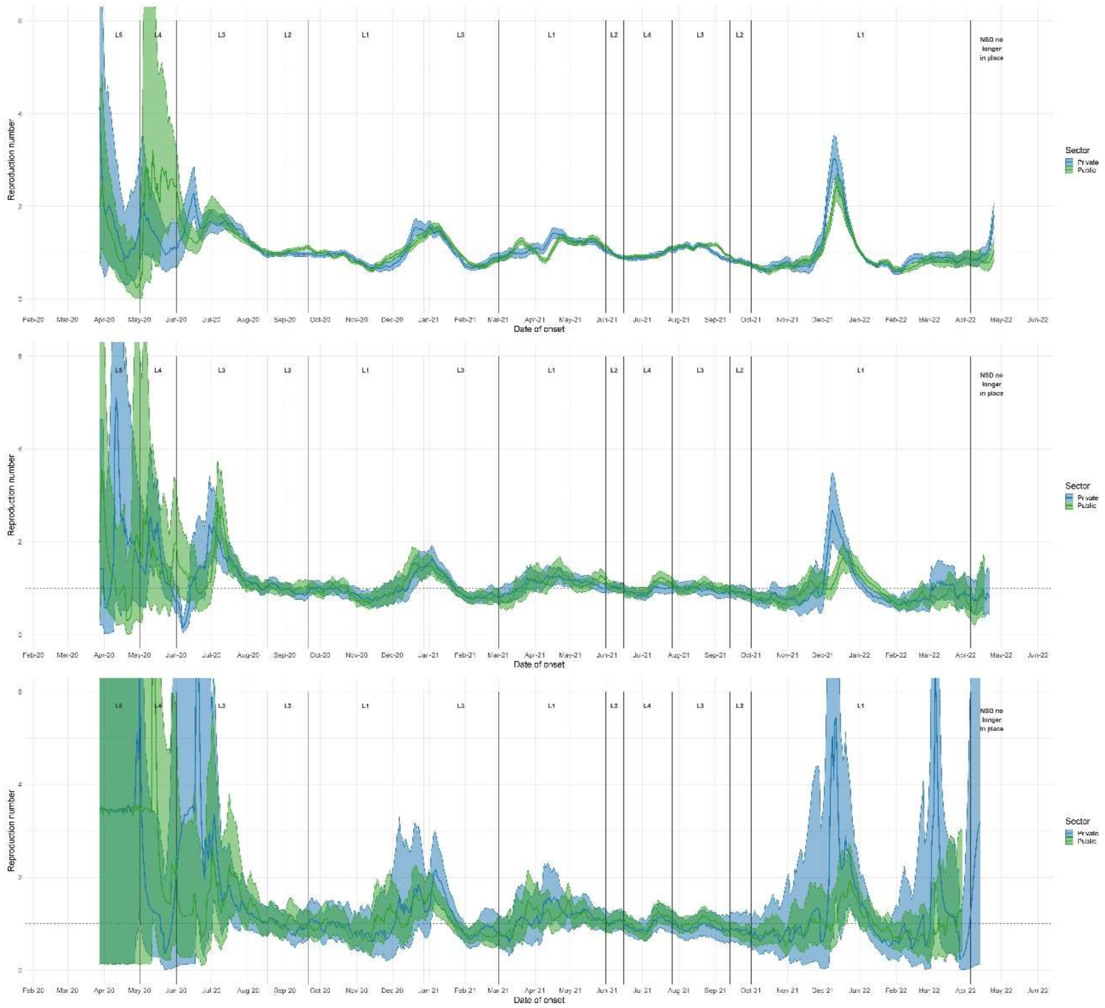


*Figure 10: R estimated separately for public and private sector data, based on cases (upper panel), admissions (middle panel) and deaths (lower panel), using 21-day sliding windows, Northern Cape. Plotted are median values, between imputations, with 2.5% and 97.5% credible intervals.*

g. North West

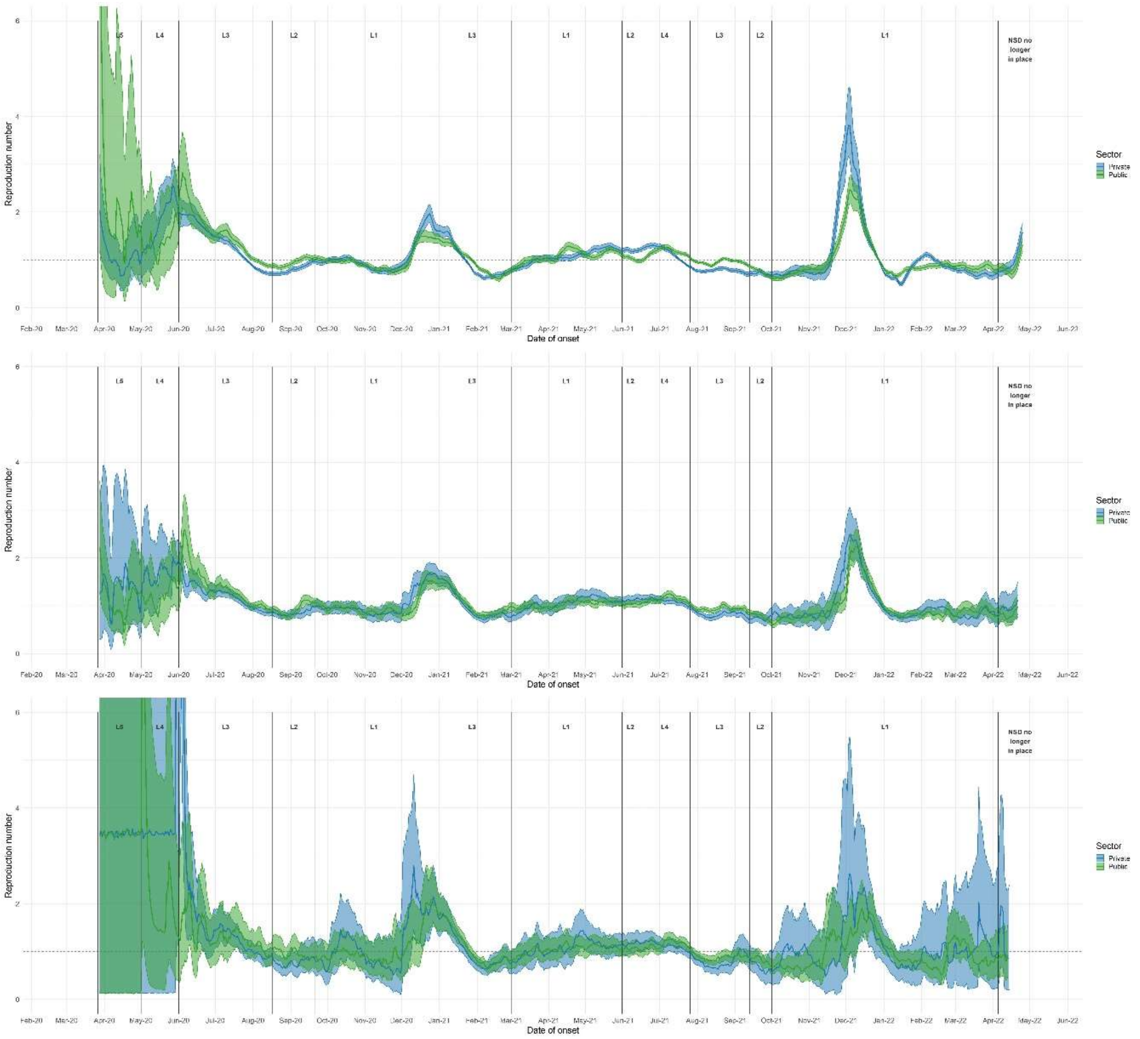


*Figure 11: R estimated separately for public and private sector data, based on cases (upper panel), admissions (middle panel) and deaths (lower panel), using 21-day sliding windows, North West. Plotted are median values, between imputations, with 2.5% and 97.5% credible intervals.*

#### h. Western Cape

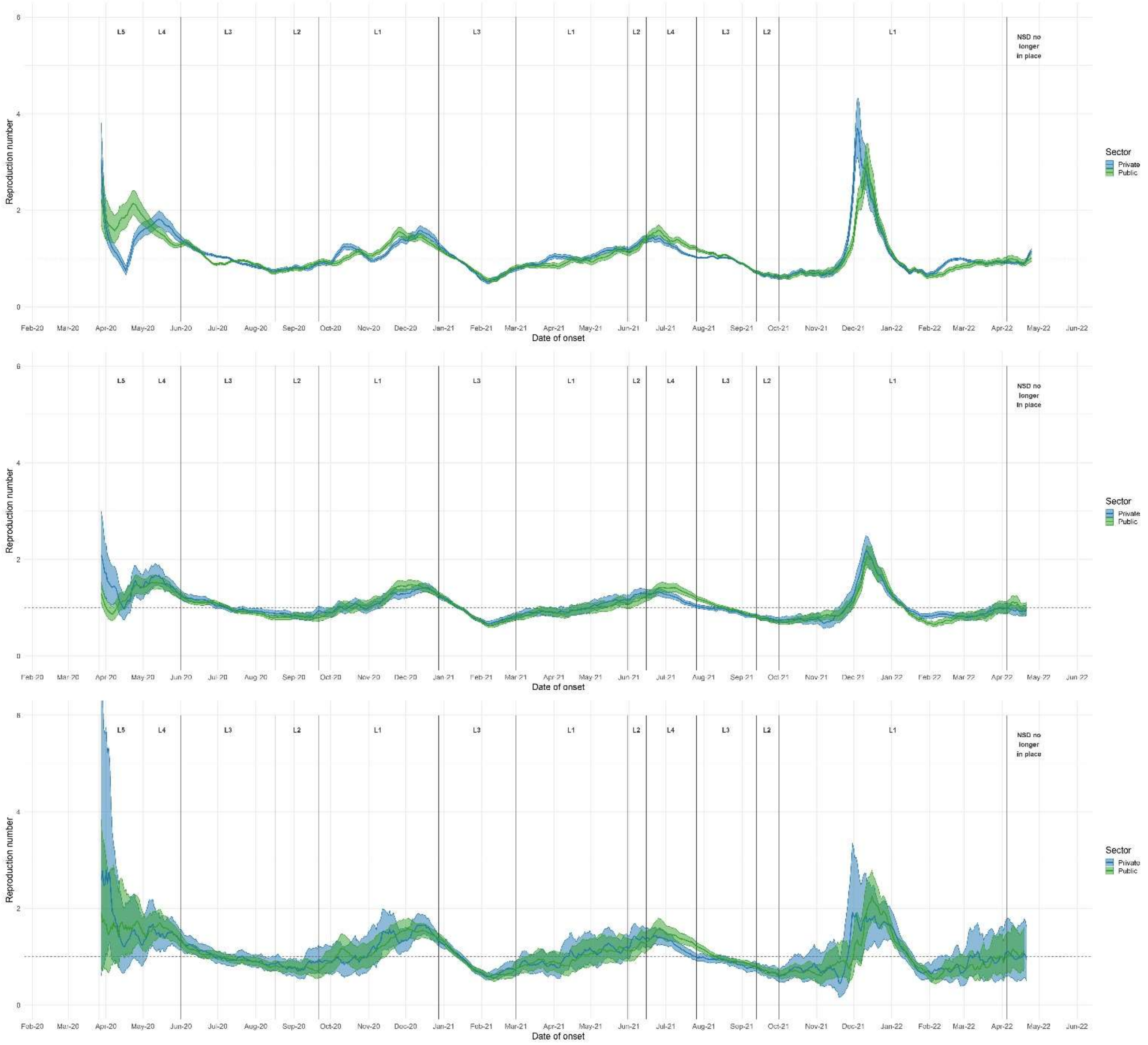


*Figure 12: R estimated separately for public and private sector data, based on cases (upper panel), admissions (middle panel) and deaths (lower panel), using 21-day sliding windows, Western Cape. Plotted are median values, between imputations, with 2.5% and 97.5% credible intervals.*

### 4. Maximum and minimum R estimates for the first four waves

| Province | Max $R_{cases}$ | | | | Max $R_{Admissions}$ | | | | Max $R_{Deaths}$ | | | |
|---|---|---|---|---|---|---|---|---|---|---|---|---|
| | Wave 1 | Wave 2 | Wave 3 | Wave 4 | Wave 1 | Wave 2 | Wave 3 | Wave 4 | Wave 1 | Wave 2 | Wave 3 | Wave 4 |
| National | 1.55 (1.43, 1.66) | 1.56 (1.47, 1.64) | 1.46 (1.38, 1.53) | 3.33 (2.83, 3.97) | 1.44 (1.37, 1.63) | 1.43 (1.37, 1.51) | 1.27 (1.23, 1.32) | 2.31 (2.08, 2.56) | 1.83 (1.44, 2.48) | 1.48 (1.38, 1.57) | 1.33 (1.26, 1.39) | 2.11 (1.83, 2.4) |
| Eastern Cape | 2.59 (1.9, 3.5) | 1.56 (1.45, 1.66) | 1.59 (1.48, 1.74) | 4.04 (3.35, 4.88) | 1.9 (1.3, 2.9) | 1.67 (1.48, 1.86) | 1.55 (1.28, 2.03) | 3.03 (2.49, 3.69) | 2.68 (1.57, 5.16) | 1.82 (1.45, 2.31) | 1.76 (1.31, 2.85) | 3.53 (2.5, 4.9) |
| Free State | 2.34 (1.82, 3.2) | 1.86 (1.7, 2.03) | 1.41 (1.31, 1.51) | 4.07 (3.33, 4.89) | 2.27 (1.63, 4.15) | 1.92 (1.67, 2.19) | 1.31 (1.19, 1.49) | 2.84 (2.37, 3.41) | 8.31 (1.61, 27.63) | 2.04 (1.6, 2.92) | 1.53 (1.13, 1.99) | 2.8 (1.86, 4.06) |
| Gauteng | 2.08 (1.87, 2.3) | 2.11 (1.89, 2.32) | 1.59 (1.5, 1.7) | 3.29 (2.77, 3.92) | 1.8 (1.64, 2.23) | 1.63 (1.5, 1.76) | 1.36 (1.29, 1.43) | 2.37 (2.08, 2.69) | 2.85 (1.75, 4.81) | 1.93 (1.63, 2.37) | 1.39 (1.29, 1.53) | 2.63 (1.95, 3.49) |
| KwaZulu-Natal | 2.03 (1.84, 2.23) | 2.2 (1.98, 2.46) | 1.72 (1.59, 1.84) | 4.37 (3.57, 5.33) | 2.54 (2.21, 2.89) | 1.83 (1.65, 2.02) | 1.71 (1.53, 1.91) | 3.16 (2.66, 3.67) | 2.26 (1.59, 3.83) | 2.02 (1.76, 2.47) | 1.7 (1.47, 2.2) | 2.57 (1.94, 3.45) |
| Limpopo | 2.61 (1.76, 4.48) | 2.16 (1.94, 2.42) | 1.85 (1.7, 2.03) | 3.66 (3.03, 4.48) | 2.41 (1.38, 5.3) | 2.09 (1.8, 2.45) | 1.57 (1.41, 1.78) | 3.62 (2.94, 4.42) | 8.43 (1.58, 27.98) | 2.67 (1.86, 4.22) | 1.96 (1.53, 2.98) | 2.68 (1.86, 3.92) |
| Mpumalanga | 2.92 (1.75, 4.81) | 1.88 (1.72, 2.05) | 1.52 (1.44, 1.62) | 3.95 (3.27, 4.74) | 6.56 (2.42, 21.18) | 1.95 (1.67, 2.25) | 1.4 (1.28, 1.53) | 2.74 (2.27, 3.33) | 8.1 (2.29, 25.69) | 2.82 (2.19, 3.59) | 1.46 (1.18, 1.85) | 3.42 (1.82, 7.09) |
| Northern Cape | 3.52 (1.73, 9.03) | 1.58 (1.47, 1.7) | 1.68 (1.53, 1.82) | 2.82 (2.43, 3.27) | 3.25 (2.21, 6.87) | 1.94 (1.54, 2.38) | 1.45 (1.21, 1.8) | 2.3 (1.8, 2.85) | 7.05 (1.86, 19.83) | 2.55 (1.68, 4.16) | 1.75 (1.24, 2.81) | 3.15 (1.54, 5.85) |
| North West | 2.82 (2.27, 4.04) | 1.88 (1.69, 2.08) | 1.36 (1.29, 1.43) | 3.53 (2.93, 4.12) | 2.26 (1.69, 4.03) | 1.73 (1.45, 2.05) | 1.28 (1.2, 1.37) | 3.29 (2.71, 3.9) | 12.81 (3.73, 31.75) | 2.91 (1.78, 4.84) | 1.41 (1.21, 1.9) | 3.17 (1.91, 4.94) |
| Western Cape | 2.2 (1.93, 2.51) | 1.58 (1.5, 1.68) | 1.58 (1.49, 1.68) | 4.13 (3.39, 5.03) | 1.77 (1.49, 2.07) | 1.49 (1.4, 1.61) | 1.46 (1.35, 1.56) | 2.47 (2.18, 2.86) | 1.86 (1.47, 3.22) | 1.64 (1.45, 1.93) | 1.62 (1.42, 1.88) | 2.38 (1.83, 3.6) |

#### 5. Choosing suitable numbers of imputations, serial interval distribution samples, and R estimates: managing computational costs of imputation and R estimation procedures

The imputation procedure was the most computationally expensive step in our analysis pipeline. As such, we wished to identify a number of imputations to conduct which would ensure consistent results while allowing analyses to be produced timeously. To this end we ran our analysis using 5, 25, 50, and 100 imputations, and compared resulting estimates (figure 18 below). Based on these results we decided to use 25 imputations, as this is cheap enough to produce timeous estimates, although the consistency with results based on 5 imputations suggests that fewer may also be sufficient.

The R estimation procedure used in this work incorporates uncertainty in the serial interval distribution through a 2-step sampling procedure: $n_1$ pairs of mean and standard deviation values are sampled, each defining a serial interval distribution; for each of the $n_1$ pairs, $n_2$ sets of R estimates are produced (for additional details see the supplementary materials in Cori et al., 2013). As such, the basic R estimation procedure is performed a total of $n_1 \times n_2$ times for each imputation. The default parameters for the EpiEstim package are $n_1 = n_2 = 1000$, leading to $1{,}000{,}000$ R estimation procedures per imputation, with significant computational cost. As such we wished to explore whether smaller choices for $n_1$ and $n_2$ would impact results, and if possible to identify parameter values which would allow cheaper R estimation without compromising the reliability or replicability of the resulting estimates. To this end we generated and compared estimates using $n_1$ values of 5, 25, 50, and 100, 500, and 1000, with $n_1 = n_2$ (figure 19). Based on these results, we settled on $n_1 = n_2 = 25$, as this was cheap enough for our purposes, and results in R estimates extremely similar to those using $n_1 = n_2 = 100$ (16x the computational expense of $n_1 = n_2 = 25$).

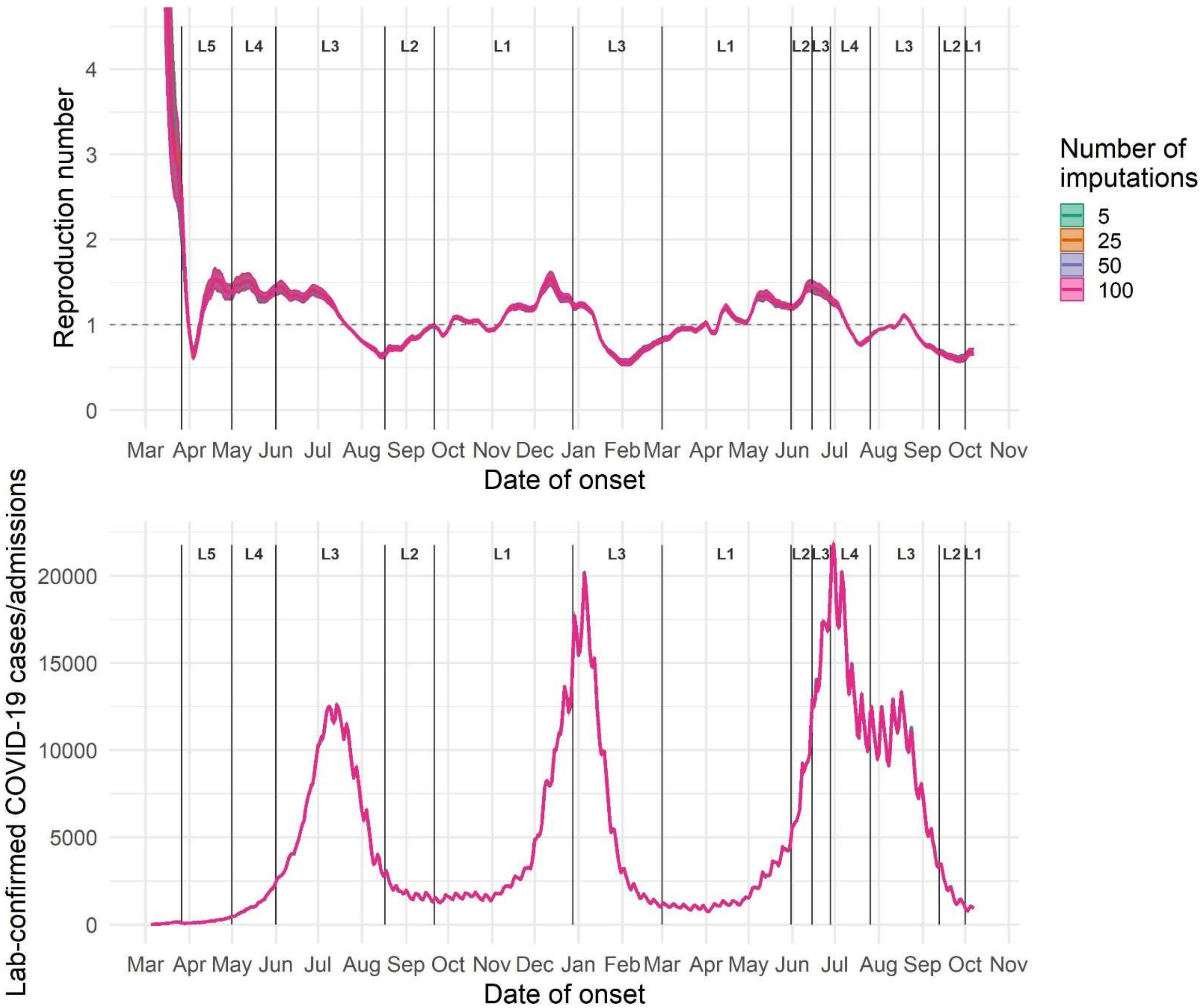


*Figure 13 R estimates (above) and imputed time series (below) using data on laboratory-confirmed infections, with a 7-day sliding window, for 5, 25, 50, and 100 imputations.*

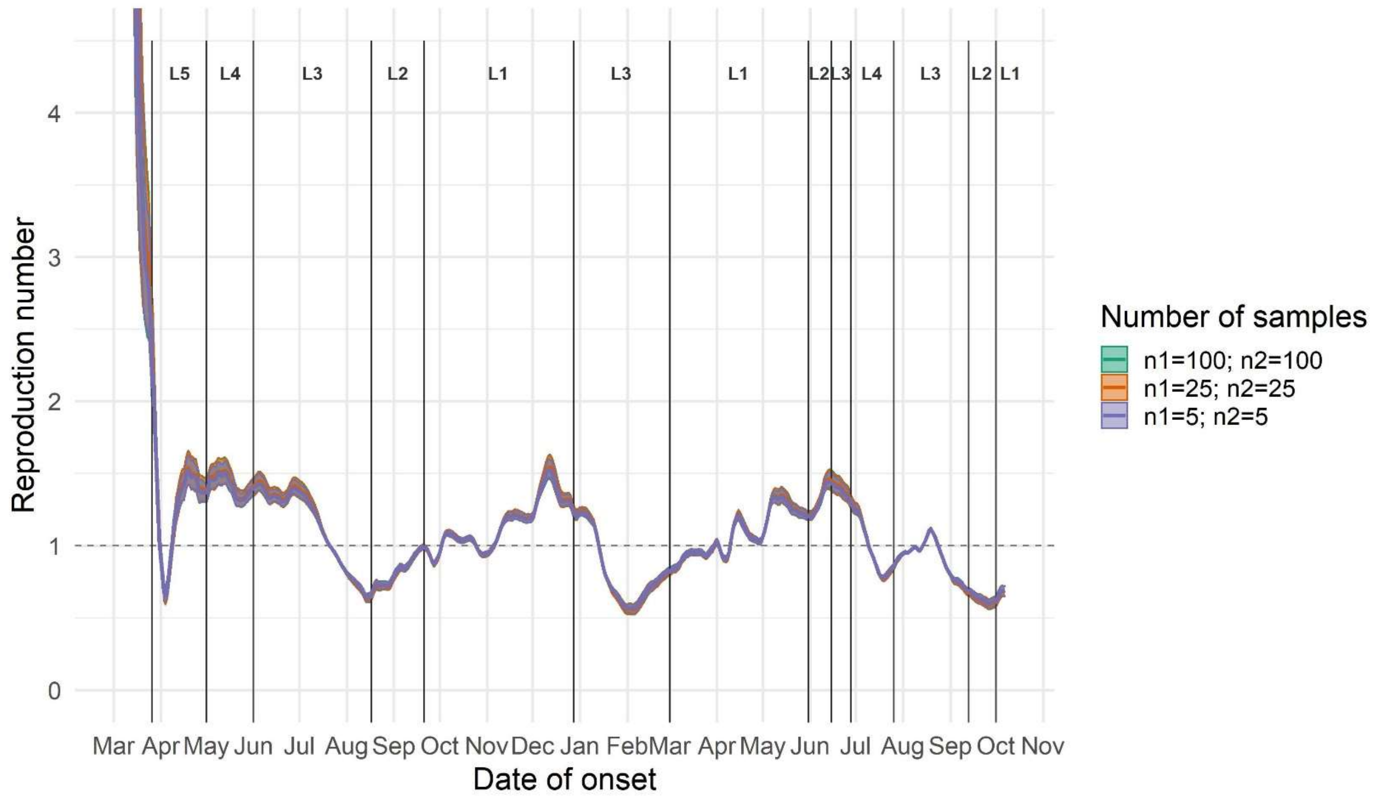


*Figure 14 R estimates based on imputed time series of laboratory-confirmed infections by dates of sypmtom onset, with a 7-day sliding window and 25 imputations, for different numbers of samples of parameter distributions (n1) and generation interval distributions (n2).*

#### 6. Ratios between data endpoints

Suitable time series data for R estimation should represent a constant proportion of true underlying incidence. To explore the validity of this assumption for our three data endpoints (laboratory-confirmed cases, hospital admissions, and hospital-associated deaths), we plot ratios of the 21-day rolling mean total hospital admissions and deaths to the 21-day rolling mean total laboratory-confirmed cases. Ratios vary substantially over the course of the epidemic, suggesting that similarities between R estimates based on the different data endpoints may be due in part to non-overlapping biases acting in opposite directions.

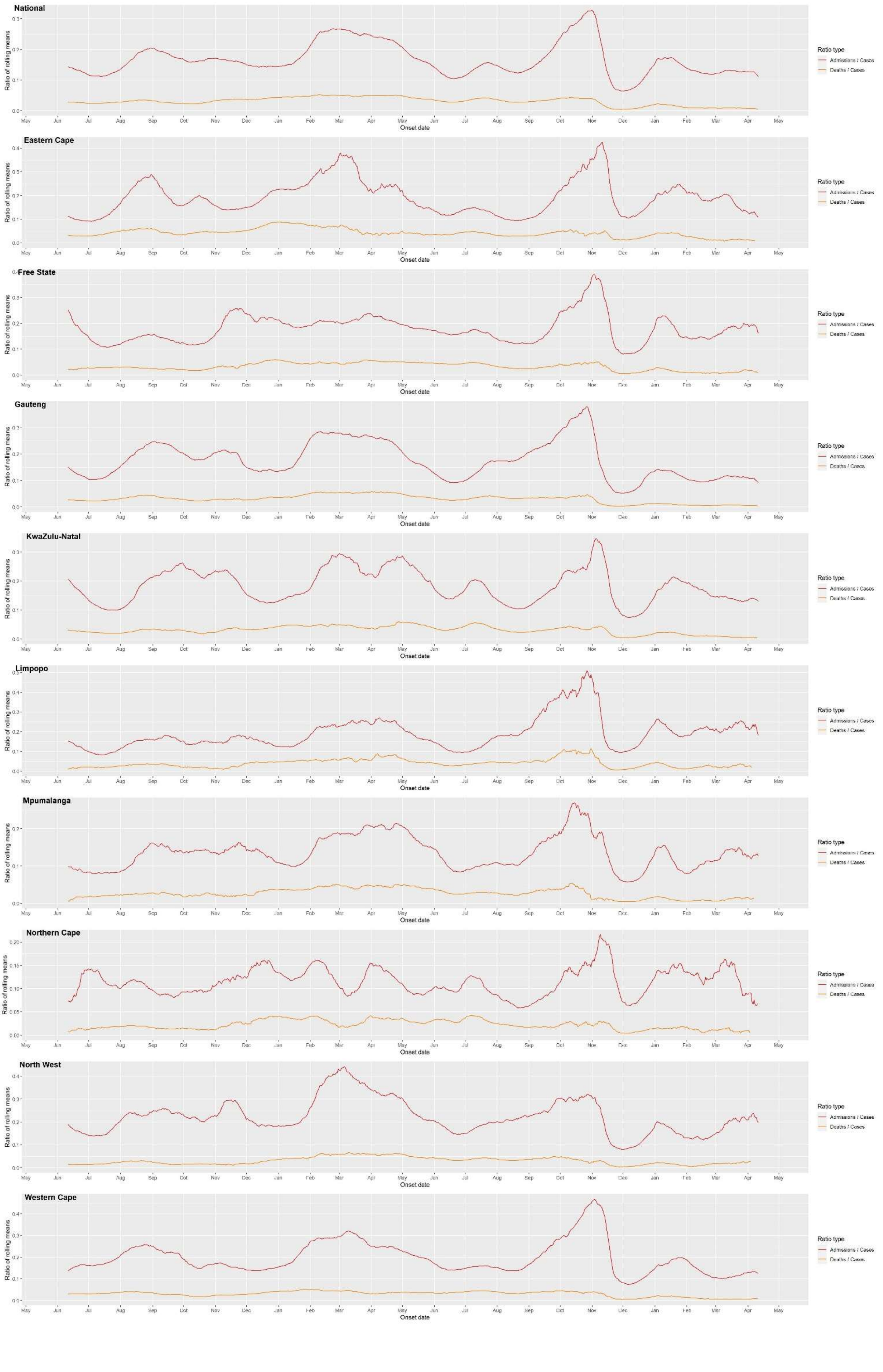
National
Eastern Cape
Free State
Gauteng
KwaZulu-Natal
Limpopo
Mpumalanga
Northern Cape
North West
Western Cape
Ratio of rolling means
Onset date
Ratio type
Admissions / Cases
Deaths / Cases

#### 7. Tests conducted per sector

Data on all COVID-19 PCR and antigen tests conducted in South African laboratories are collected by the NICD. The dataset consists of 16 729 736 tests conducted between 1 March 2020 and 6 September 2021. We plot metrics on testing programs by “specimen received date” – the date on which a testing laboratory received each sample – as this column is complete (specimen collection dates are missing 18340 values and test result reporting dates are missing 1102 values).

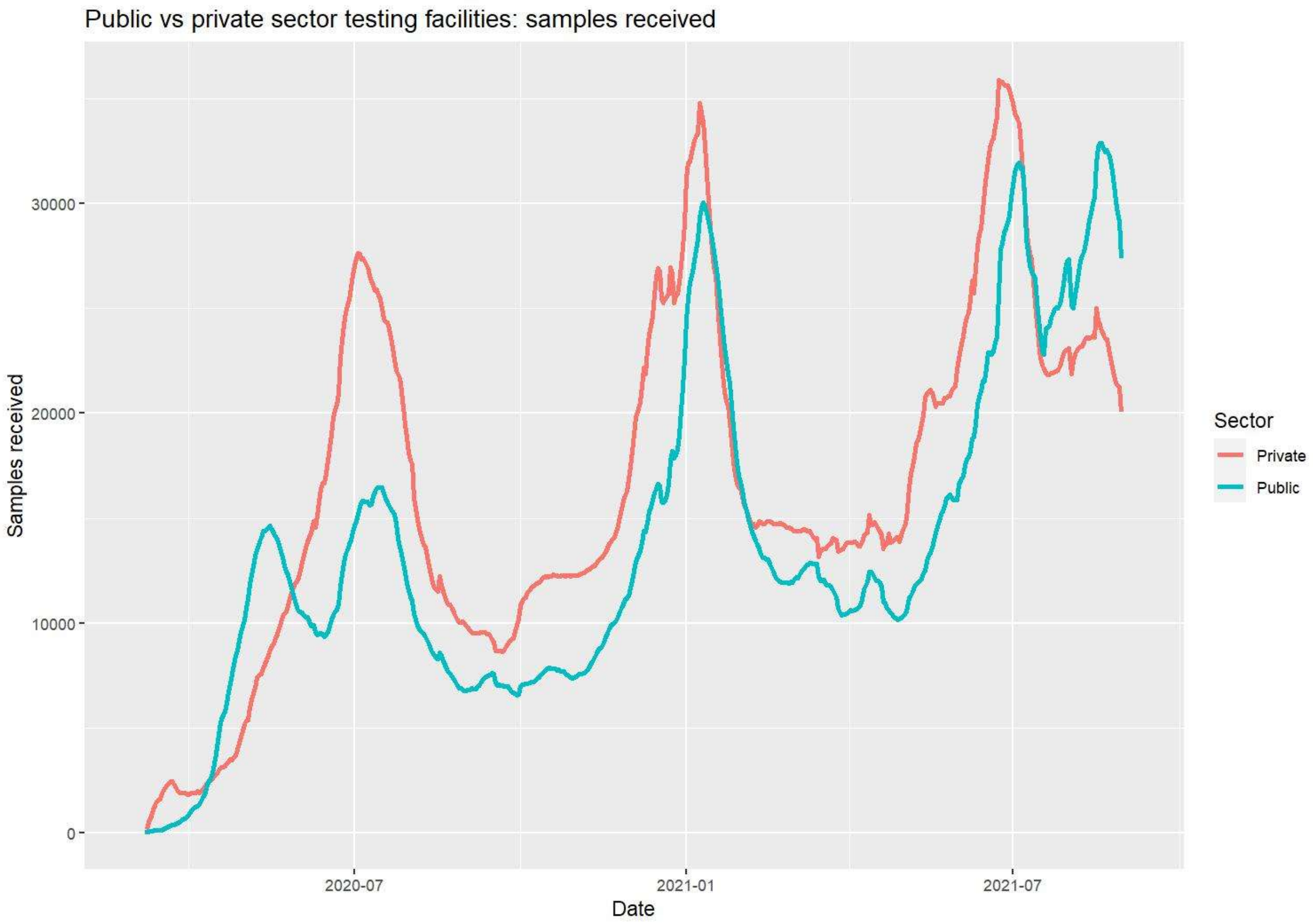

Public vs private sector testing facilities: samples received
Samples received
30000
20000
10000
0
2020-07
2021-01
2021-07
Date
Sector
Private
Public

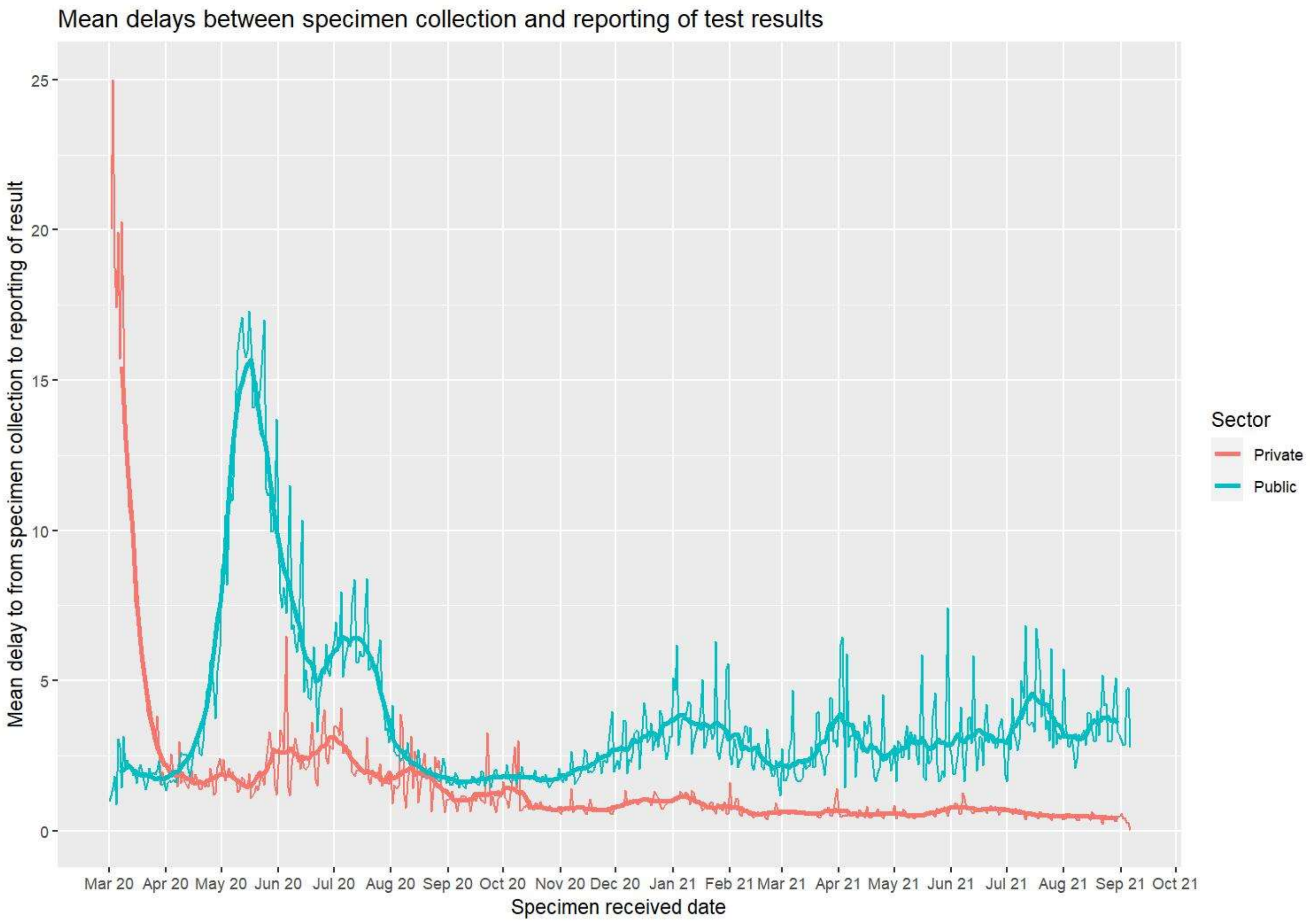
Mean delays between specimen collection and reporting of test results
Mean delay to from specimen collection to reporting of result
25
20
15
10
5
0
Mar 20
Apr 20
May 20
Jun 20
Jul 20
Aug 20
Sep 20
Oct 20
Nov 20
Dec 20
Jan 21
Feb 21
Mar 21
Apr 21
May 21
Jun 21
Jul 21
Aug 21
Sep 21
Oct 21
Specimen received date
Sector
Private
Public

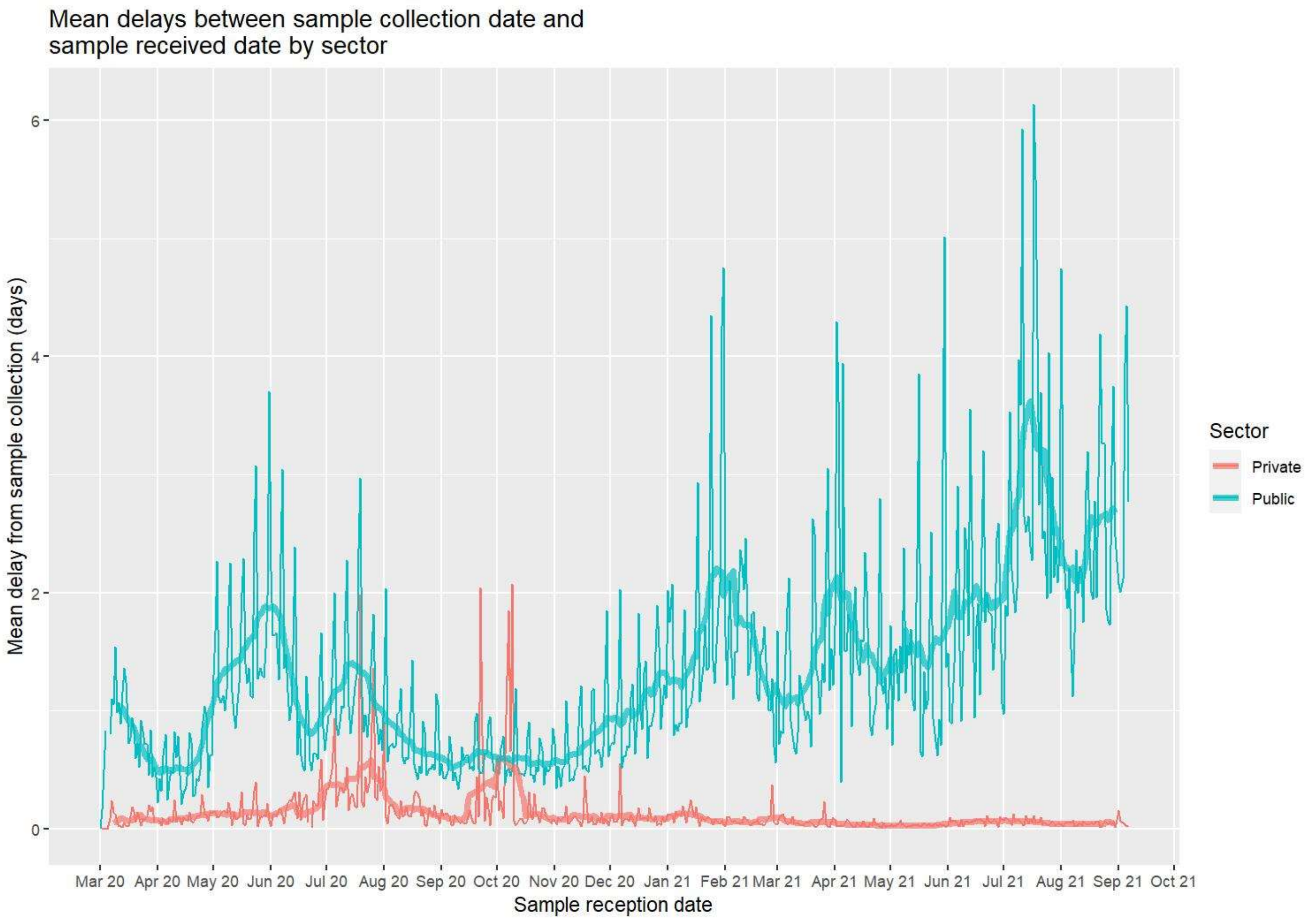
Mean delays between sample collection date and
sample received date by sector
Mean delay from sample collection (days)
0
2
4
6
Mar 20
Apr 20
May 20
Jun 20
Jul 20
Aug 20
Sep 20
Oct 20
Nov 20
Dec 20
Jan 21
Feb 21
Mar 21
Apr 21
May 21
Jun 21
Jul 21
Aug 21
Sep 21
Oct 21
Sample reception date
Sector
Private
Public
